# Familial and Parental Relationships Can Ease Anxiety Among Academics in the United States

**DOI:** 10.1101/2025.04.30.25326730

**Authors:** Anietie Andy, Natasha Tonge, Praise-EL Michaels, Marina K. Holz

**Author notes:** Correspondence concerning this article should be addressed to Anietie Andy, Electrical Engineering and Computer Science, Howard University, 2300 6th St NW, Washington, DC.

## Abstract

Anxiety among academic faculty remains a critical yet understudied public health concern because of comorbidity with depression and other mental health disorders. Here, we present a large study examining faculty anxiety, analyzing data from 2,106 professors across 62 U.S. higher education institutions using the Generalized Anxiety Disorder (GAD-7) assessment. This comprehensive dataset revealed previously unknown patterns in how familial relationships and academic lineage influence faculty anxiety. The analysis in this work focused on faculty members who are not in a health profession, The exclusion of health profession faculty members in this study is important because compared to non-health profession faculty members, they experience a distinct set of mental health challenges which are exacerbated by clinical practice. Our analysis uncovered a clear relationship between academic rank and anxiety levels, with tenure-track assistant professors showing the highest anxiety scores (predicted GAD-7 = 7.9) compared to full professors. Although academic discipline explains only 0.32% of anxiety variance, we identify institutional factors and family support as major moderators of faculty mental health. We find that close family relationships significantly moderated anxiety across institution types, with particularly strong protective effects at Historically Black Colleges and Universities (HBCUs) / Hispanic Serving Institutions (HSIs), b = −4.24, p = .015 and public institutions, b = −1.11, p = .027. These findings address a crucial gap in understanding faculty mental health and suggest that institutions should implement targeted interventions focusing on reinforcing social support systems, particularly for early-career academics.

## Introduction

Anxiety is a mood state characterized by fear or worry for the future and often associated with muscle tension, vigilance, and avoidance^1^. Anxiety is highly comorbid with other mental health conditions such as depression, with anxiety preceding depression in over 75% of affected individuals^2^, and increasing vulnerability to other health conditions, such as unhealthy behaviors and chronic disease^3^. Globally, anxiety presents an unaddressed mental health need and a significant public health burden^4^. Academic faculty lead complex professional lives and are uniquely susceptible to anxiety-inducing factors. Faculty members integrate multiple demanding responsibilities across scholarly research, teaching, and administrative duties, leading to chronic role overload and significant time management challenges^5^. Career progression requirements on competitive promotion and tenure track add a layer of instability and uncertainty. Academic success hinges on securing research funding, publish-or-perish academic culture, producing high-impact scholarly output and navigating rigorous peer review processes, further exacerbating anxiety. Financial pressures add another layer of stress, including relatively low compensation compared to career trajectories outside the academy, and ongoing student loan burden. The hierarchical institutional structures, combined with cultural expectations of persistent high performance, create an environment where mental health challenges are marginalized.

Several studies have investigated mental health and anxiety expression in undergraduate, medical and doctoral students^6–10^. In contrast, while some attention has been dedicated to concerns of mental health among students and faculty^11,27--33^, few studies have systematically analyzed anxiety in academic faculty. A study of non-tenure track faculty found that contingent faculty with lower family incomes, inability to find permanent positions, high institutional commitment, and use of dysfunctional coping mechanisms were more likely to experience depression, anxiety, and stress^12^. A single-university study of students and faculty/staff found that approximately one-third of both groups experienced severe or extremely severe symptoms of depression, anxiety, or stress^13^, and others have observed that the experience of psychological distress varied with career stage in postgraduate students, early-middle career researchers and senior researchers^14^. Prior work ^29^ executed a national study to assess occupational stress among academic staff in 56 universities in Canada. It was found that stressors such as work-life conflict and job security impacted the outcome of health and workplace satisfaction. A study surveyed 5,500 participants from a university and its academic medical center to assess the mental health and well-being of their clinical and non-clinical workers during the early phase of the COVID-19 pandemic. It was found that clinical or community COVID-19 exposure, poor supervisor support, people who were 40 years and older, and family/home stressors were linked to worse health outcomes ^30^. Existing literature also highlights the prevalence of mental health concerns among university faculty, identifying factors such as workload, work-life balance issues, job insecurity, and pressure to perform as contributors to mental health issues^15^. Thus, previous findings emphasize the need for comprehensive analysis of the prevalence of anxiety in academic professors to determine the need for institutional interventions that target anxiety to improve faculty health well-being. Nonetheless, there is a lack of understanding of the general prevalence of anxiety among postsecondary faculty, and the impact of modifiers on this distinct mental health challenge. Therefore, there exists a critical need to study the expression of anxiety among academic faculty, to inform tailored interventions that can take into account both individual resilience and systemic institutional factors and create durable and meaningful support systems. To this end, we explored how close familial relationships and having an academic parent or partner modifies the experience of anxiety in academics from different institution types, academic disciplines and ranks.

## Methods

### Measures

In order to systematically assess the prevalence of anxiety among academic faculty members in the United States, we administered the Generalized Anxiety Disorder (GAD-7) questionnaire [16] as well as a demographic questionnaire to academic faculty from 62 higher education and medical institutions such as Liberal Arts Colleges, Historically Black Colleges and Universities (HBCU) or Hispanic Serving Institutions (HSI), Medical Schools and Academic Medical Centers, and Community Colleges.. The GAD-7 is a well-validated seven-item instrument where scores of 5-9 indicate mild anxiety, 10-14 indicate moderate anxiety, and 15-21 indicates severe anxiety^16^. In addition to demographics, we asked about close family relationships, presence of an academic parent, the others were administered in a single survey in qualtrics. Close family relationships, academic parent were administered as binary responses (yes/no) and both were left open to individual interpretation. We also provided the participants with an optional text box to express their feedback about the survey or thoughts about anxiety in academia in general. The survey/questionnaire was approved by the Howard University Institutional Review Board. Data was collected using Qualtrics; descriptive statistics and graphs were made using R^17^. Informed consent was obtained from all participants. The informed consent was shared with participants via email. Due to the IRB stipulations, the collected data used for this study cannot be shared or made publicly accessible.,

### Participants

The survey was administered over a period of 6 months i.e. from June to December 2024. The survey was designed to be anonymous, meaning that there is no way to connect participants’ responses to the participants. The survey was sent to the academic email addresses of faculty members; this survey. 3,358 faculty members responded to the survey. We then identified participants with a survey completion time greater than 3 standard deviations above the mean completion time of all the participants to remove potential survey outliers who may have been distracted or inattentive^18^. 2,655 participants met these criteria. Out of these, we excluded 549 participants who identified as having a health profession specialty (e.g., medicine, dentistry, pharmacy, physical therapy), for a final participant number of 2,106. In this work, we focus on analyzing data by faculty members who are not in a health profession because health professions faculty experience a distinct set of mental health challenges exacerbated by clinical practice^19^, and further exacerbated by the COVID-19 pandemic^20^. Table 1 depicts demographic characteristics of the survey participants.

**Table 1.**
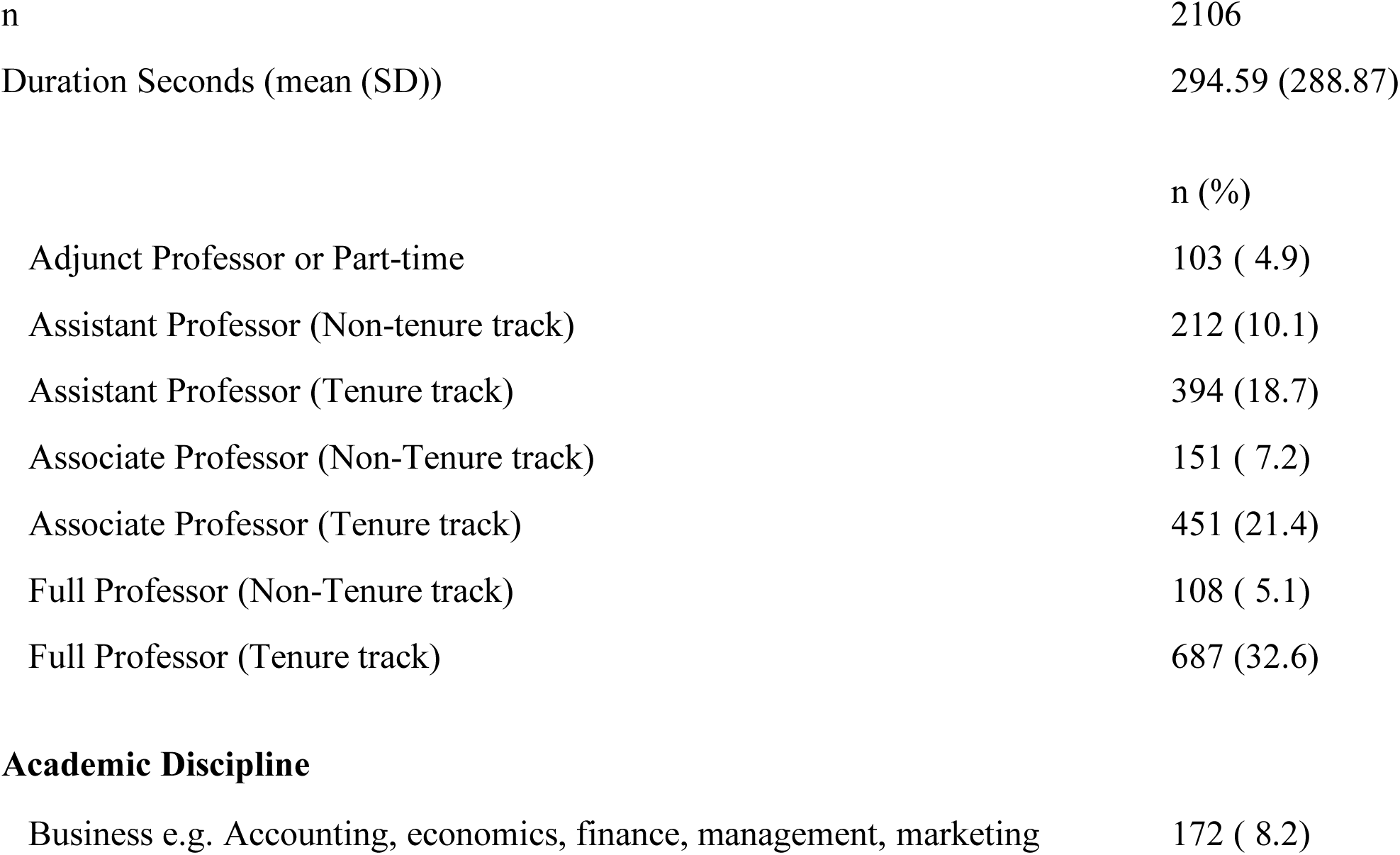
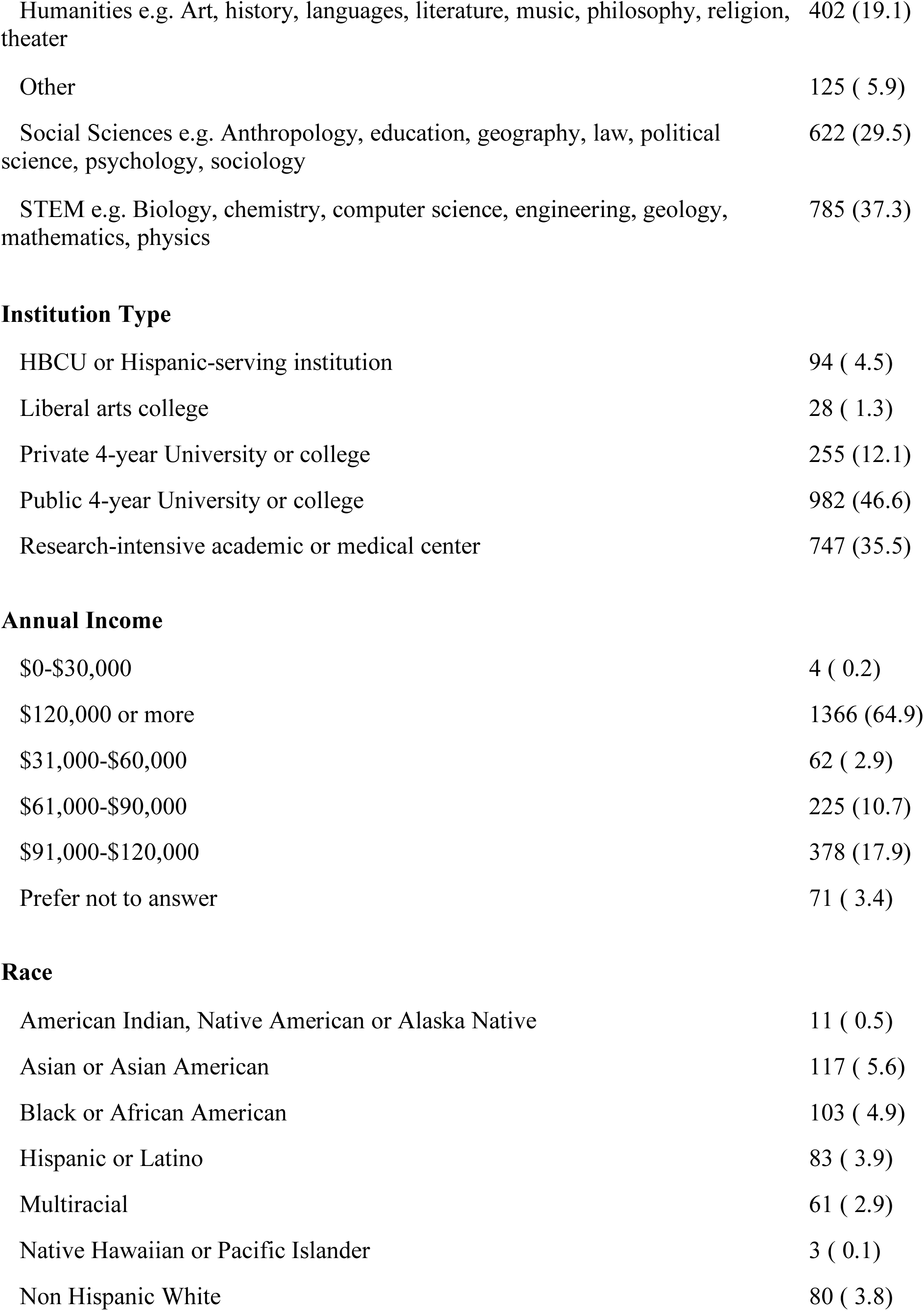
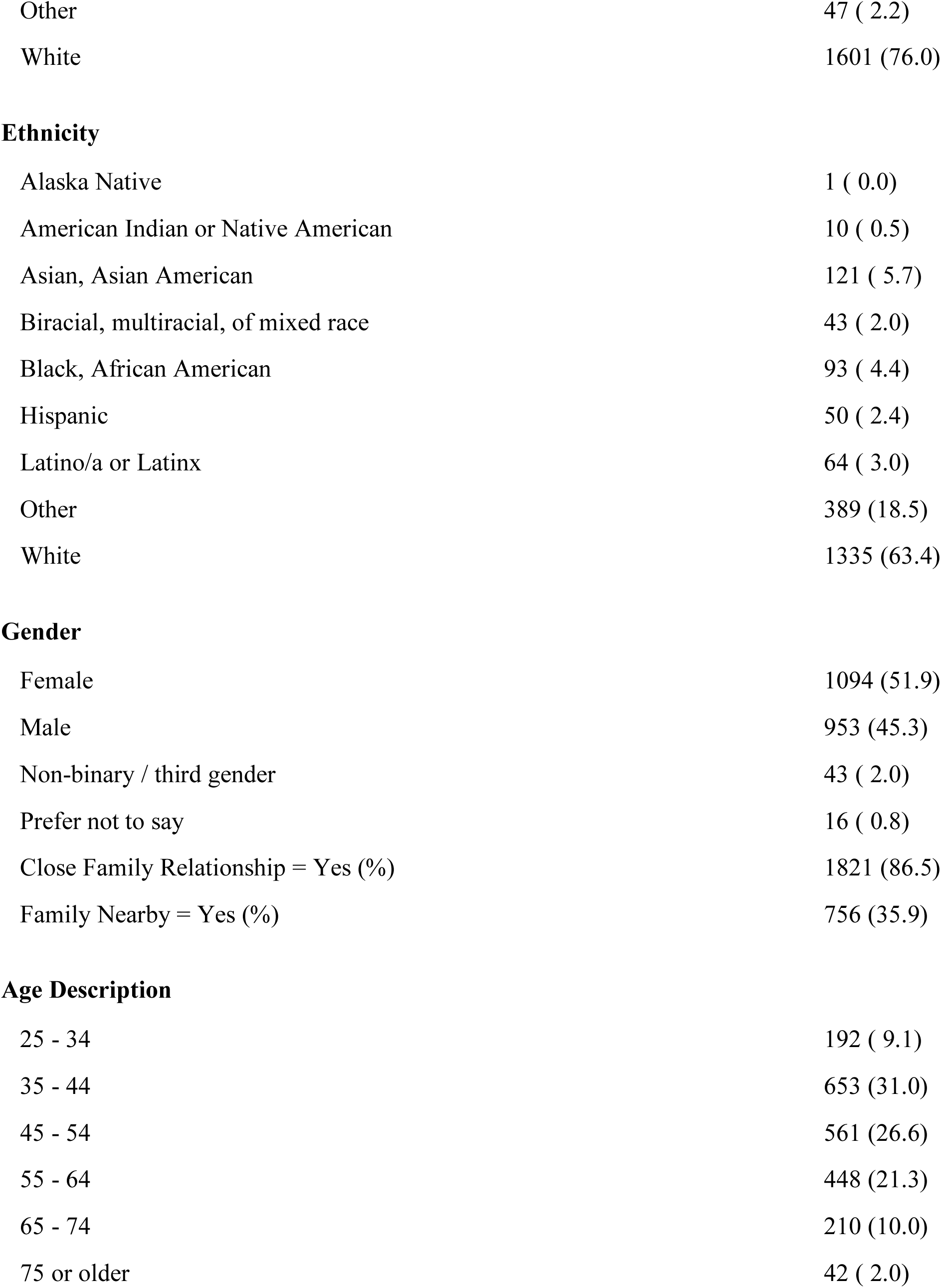
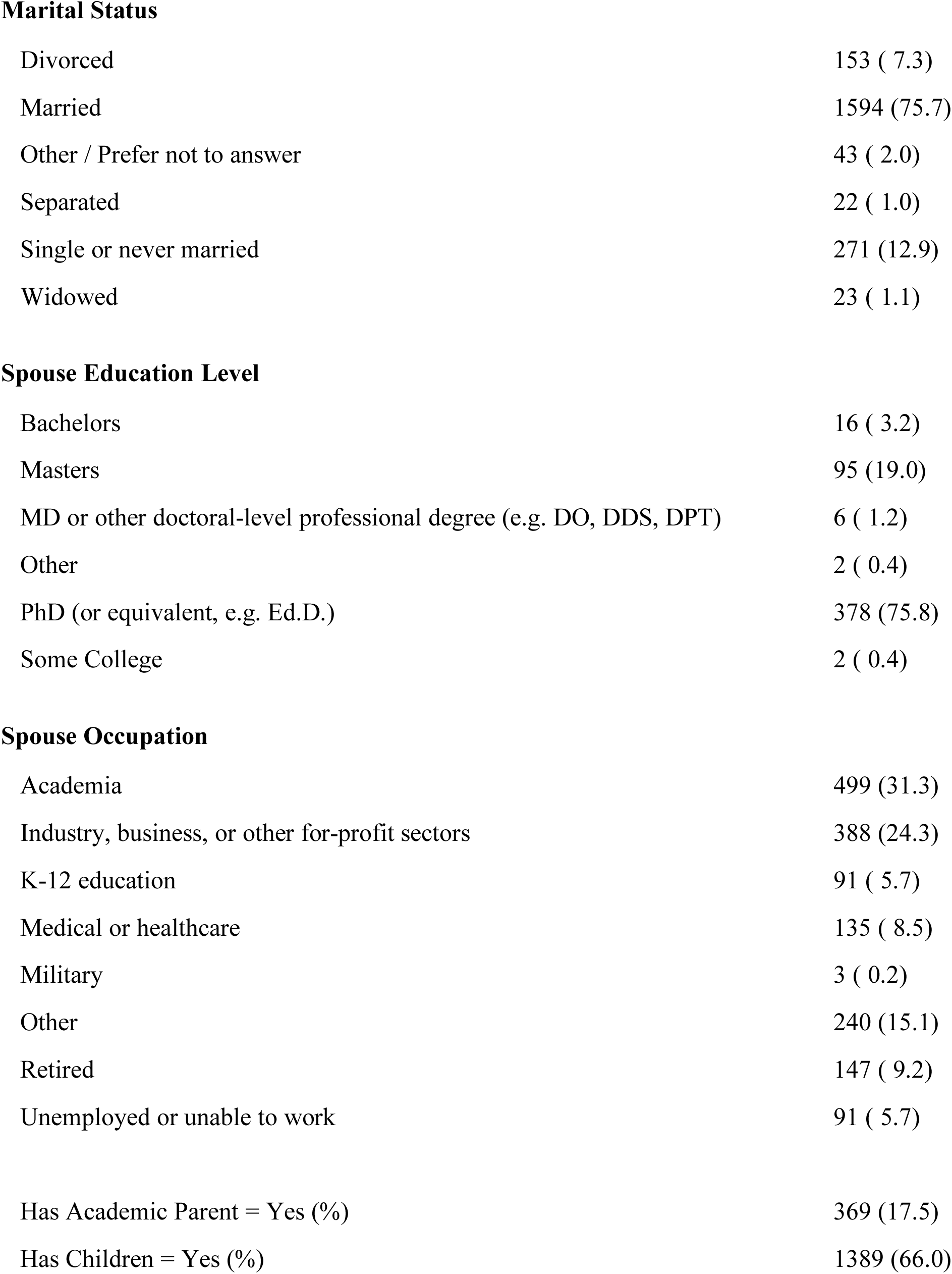
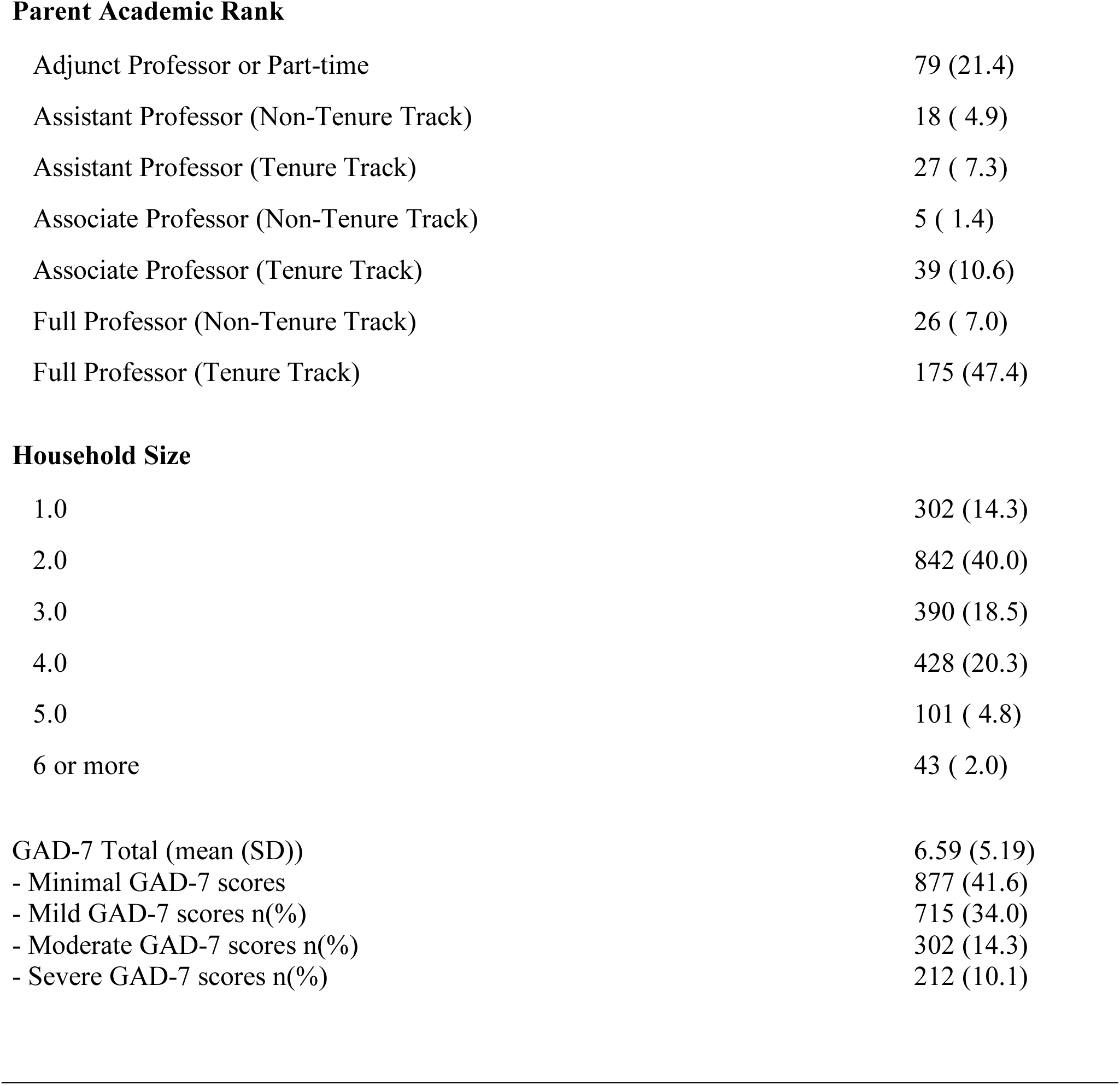
Respondent Demographics and Characteristics.

|  |  |
| --- | --- |
| n | 2106 |
| Duration Seconds (mean (SD)) | 294.59 (288.87) |
|  | n (%) |
| Adjunct Professor or Part-time | 103 ( 4.9) |
| Assistant Professor (Non-tenure track) | 212 (10.1) |
| Assistant Professor (Tenure track) | 394 (18.7) |
| Associate Professor (Non-Tenure track) | 151 ( 7.2) |
| Associate Professor (Tenure track) | 451 (21.4) |
| Full Professor (Non-Tenure track) | 108 ( 5.1) |
| Full Professor (Tenure track) | 687 (32.6) |
| **Academic Discipline** |  |
| Business e.g. Accounting, economics, finance, management, marketing | 172 ( 8.2) |
| Humanities e.g. Art, history, languages, literature, music, philosophy, religion, theater | 402 (19.1) |
| Other | 125 ( 5.9) |
| Social Sciences e.g. Anthropology, education, geography, law, political science, psychology, sociology | 622 (29.5) |
| STEM e.g. Biology, chemistry, computer science, engineering, geology, mathematics, physics | 785 (37.3) |
| **Institution Type** |  |
| HBCU or Hispanic-serving institution | 94 ( 4.5) |
| Liberal arts college | 28 ( 1.3) |
| Private 4-year University or college | 255 (12.1) |
| Public 4-year University or college | 982 (46.6) |
| Research-intensive academic or medical center | 747 (35.5) |
| **Annual Income** |  |
| \$0-\$30,000 | 4 ( 0.2) |
| \$120,000 or more | 1366 (64.9) |
| \$31,000-\$60,000 | 62 ( 2.9) |
| \$61,000-\$90,000 | 225 (10.7) |
| \$91,000-\$120,000 | 378 (17.9) |
| Prefer not to answer | 71 ( 3.4) |
| **Race** |  |
| American Indian, Native American or Alaska Native | 11 ( 0.5) |
| Asian or Asian American | 117 ( 5.6) |
| Black or African American | 103 ( 4.9) |
| Hispanic or Latino | 83 ( 3.9) |
| Multiracial | 61 ( 2.9) |
| Native Hawaiian or Pacific Islander | 3 ( 0.1) |
| Non Hispanic White | 80 ( 3.8) |
| Other | 47 ( 2.2) |
| White | 1601 (76.0) |
| **Ethnicity** |  |
| Alaska Native | 1 ( 0.0) |
| American Indian or Native American | 10 ( 0.5) |
| Asian, Asian American | 121 ( 5.7) |
| Biracial, multiracial, of mixed race | 43 ( 2.0) |
| Black, African American | 93 ( 4.4) |
| Hispanic | 50 ( 2.4) |
| Latino/a or Latinx | 64 ( 3.0) |
| Other | 389 (18.5) |
| White | 1335 (63.4) |
| **Gender** |  |
| Female | 1094 (51.9) |
| Male | 953 (45.3) |
| Non-binary / third gender | 43 ( 2.0) |
| Prefer not to say | 16 ( 0.8) |
| Close Family Relationship = Yes (%) | 1821 (86.5) |
| Family Nearby = Yes (%) | 756 (35.9) |
| **Age Description** |  |
| 25 - 34 | 192 ( 9.1) |
| 35 - 44 | 653 (31.0) |
| 45 - 54 | 561 (26.6) |
| 55 - 64 | 448 (21.3) |
| 65 - 74 | 210 (10.0) |
| 75 or older | 42 ( 2.0) |
| **Marital Status** |  |
| Divorced | 153 ( 7.3) |
| Married | 1594 (75.7) |
| Other / Prefer not to answer | 43 ( 2.0) |
| Separated | 22 ( 1.0) |
| Single or never married | 271 (12.9) |
| Widowed | 23 ( 1.1) |
| **Spouse Education Level** |  |
| Bachelors | 16 ( 3.2) |
| Masters | 95 (19.0) |
| MD or other doctoral-level professional degree (e.g. DO, DDS, DPT) | 6 ( 1.2) |
| Other | 2 ( 0.4) |
| PhD (or equivalent, e.g. Ed.D.) | 378 (75.8) |
| Some College | 2 ( 0.4) |
| **Spouse Occupation** |  |
| Academia | 499 (31.3) |
| Industry, business, or other for-profit sectors | 388 (24.3) |
| K-12 education | 91 ( 5.7) |
| Medical or healthcare | 135 ( 8.5) |
| Military | 3 ( 0.2) |
| Other | 240 (15.1) |
| Retired | 147 ( 9.2) |
| Unemployed or unable to work | 91 ( 5.7) |
| Has Academic Parent = Yes (%) | 369 (17.5) |
| Has Children = Yes (%) | 1389 (66.0) |
| **Parent Academic Rank** |  |
| Adjunct Professor or Part-time | 79 (21.4) |
| Assistant Professor (Non-Tenure Track) | 18 ( 4.9) |
| Assistant Professor (Tenure Track) | 27 ( 7.3) |
| Associate Professor (Non-Tenure Track) | 5 ( 1.4) |
| Associate Professor (Tenure Track) | 39 (10.6) |
| Full Professor (Non-Tenure Track) | 26 ( 7.0) |
| Full Professor (Tenure Track) | 175 (47.4) |
| **Household Size** |  |
| 1.0 | 302 (14.3) |
| 2.0 | 842 (40.0) |
| 3.0 | 390 (18.5) |
| 4.0 | 428 (20.3) |
| 5.0 | 101 ( 4.8) |
| 6 or more | 43 ( 2.0) |
| GAD-7 Total (mean (SD)) | 6.59 (5.19) |
| - Minimal GAD-7 scores | 877 (41.6) |
| - Mild GAD-7 scores n(%) | 715 (34.0) |
| - Moderate GAD-7 scores n(%) | 302 (14.3) |
| - Severe GAD-7 scores n(%) | 212 (10.1) |

## Results

Demographic information about the sample and descriptive statistics of responses can be found in Table 1. In Table 2, we report correlations between several variables of interest (e.g., academic rank, discipline, institution type, family relationship, and presence of an academic parent), and demographics (e.g., gender, race, ethnicity, income).

**Table 2.** Means, standard deviations, and correlations with confidence intervals.

| Variable | *M* | *SD* | 1 | 2 | 3 | 4 | 5 | 6 |
| --- | --- | --- | --- | --- | --- | --- | --- | --- |
| 1. GAD-7 Total | 6.51 | 5.11 |  |  |  |  |  |  |
| 2. Annual Income | 4.51 | 0.81 | -.13** |  |  |  |  |  |
|  |  |  | [-.17, -.08] |  |  |  |  |  |
| 3. Gender(Female ) | 0.54 | 0.50 | .11** | -.06** |  |  |  |  |
|  |  |  | [.07, .16] | [-.11, -.02] |  |  |  |  |
| 4. Age Group | 2.97 | 1.20 | -.20** | .17** | -.11** |  |  |  |
|  |  |  | [-.25, -.16] | [.13, .21] | [-.16, -.07] |  |  |  |
| 5. Academic Rank | 4.75 | 1.94 | -.13** | .38** | -.11** | .48** |  |  |
|  |  |  | [-.18, -.09] | [.34, .42] | [-.15, -.06] | [.44, .51] |  |  |
| 6. Has Close Family Relationship | 0.87 | 0.34 | -.06* | .07** | .04 | .07** | .04 |  |
|  |  |  | [-.10, -.01] | [.02, .11] | [-.00, .09] | [.02, .11] | [-.01, .08] |  |
| 7. Has Academic Parent | 0.18 | 0.38 | -.04 | .02 | -.01 | .04 | .05* | .04 |
|  |  |  | [-.09, .00] | [-.02, .07] | [-.05, .03] | [-.00, .08] | [.00, .09] | [-.00, .09] |

*Note.* *M* and *SD* are used to represent mean and standard deviation, respectively. Values in square brackets indicate the 95% confidence interval for each correlation. The confidence interval is a plausible range of population correlations that could have caused the sample correlation (Cumming, 2014). \* indicates $p < .05$ . \*\* indicates $p < .01$ .

Because we found a significant relationship between gender and anxiety, we included gender as a covariate in all further analyses. We did not include other demographic variables of age and income due the fact that these variables are conceptually overlapping with primary predictors of interest (e.g., higher rank faculty are typically older and have higher income). Gender was dummy coded with *female* as the reference group; *male* and *non-binary*, were dummy coded variables and *prefer not to say* was removed due to low response rates of <1%.

### Overall differences in institution type, discipline, and tenure status on anxiety among faculty

To allow for consistent interpretations, we used the category with the largest number of faculty surveyed as the reference group in each model. First, we examined univariable effects, controlling for gender. For the effect of institution type on anxiety (Figure 1), we found a small but significant overall model, F(6,2083) = 10.92, p < .001, η^2^ = .005 with an adjusted r-squared suggesting that collectively, about 2.8% of variance in anxiety was explained by institution and gender. We observed the effects for specific institutions and found that only faculty at HBCU/HSIs were significantly more anxious than counterparts at public 4-year institutions, *b* = 1.25, p = .025. Academic rank was significantly associated with anxiety, F(6,2083)=12.98, p < .001, η^2^ = .04 and the adjusted r-squared suggested that collectively, about 5.4% of the variance in faculty anxiety scores could be explained by rank and gender. Relative to tenure-track full professors, faculty at all ranks were significantly more anxious except for non-tenure track full professors, *p*s < .011. Tenure-track assistant professors have the highest levels of anxiety with an estimated GAD-7 score of 8.1. Finally, the model of academic discipline was significantly associated with anxiety overall, F(6,2083)=11.3, *p*<.001, η^2^ = .005; adjusted R-squared suggested 2.8% of variance in faculty anxiety is explained by discipline and gender. Relative to faculty in STEM, the discipline with the largest number of faculty, faculty in business were significantly less anxious, *b* = −1.10, *p* = .02, 95% CI[−1.95 – −0.253]. The model predicted that STEM faculty had GAD-7 scores around 7.34, which was the second highest of the disciplines following faculty in the Humanities with predicted GAD-7 scores of around 7.3. All effects can be viewed in Table 3.

**S Figure 1.**
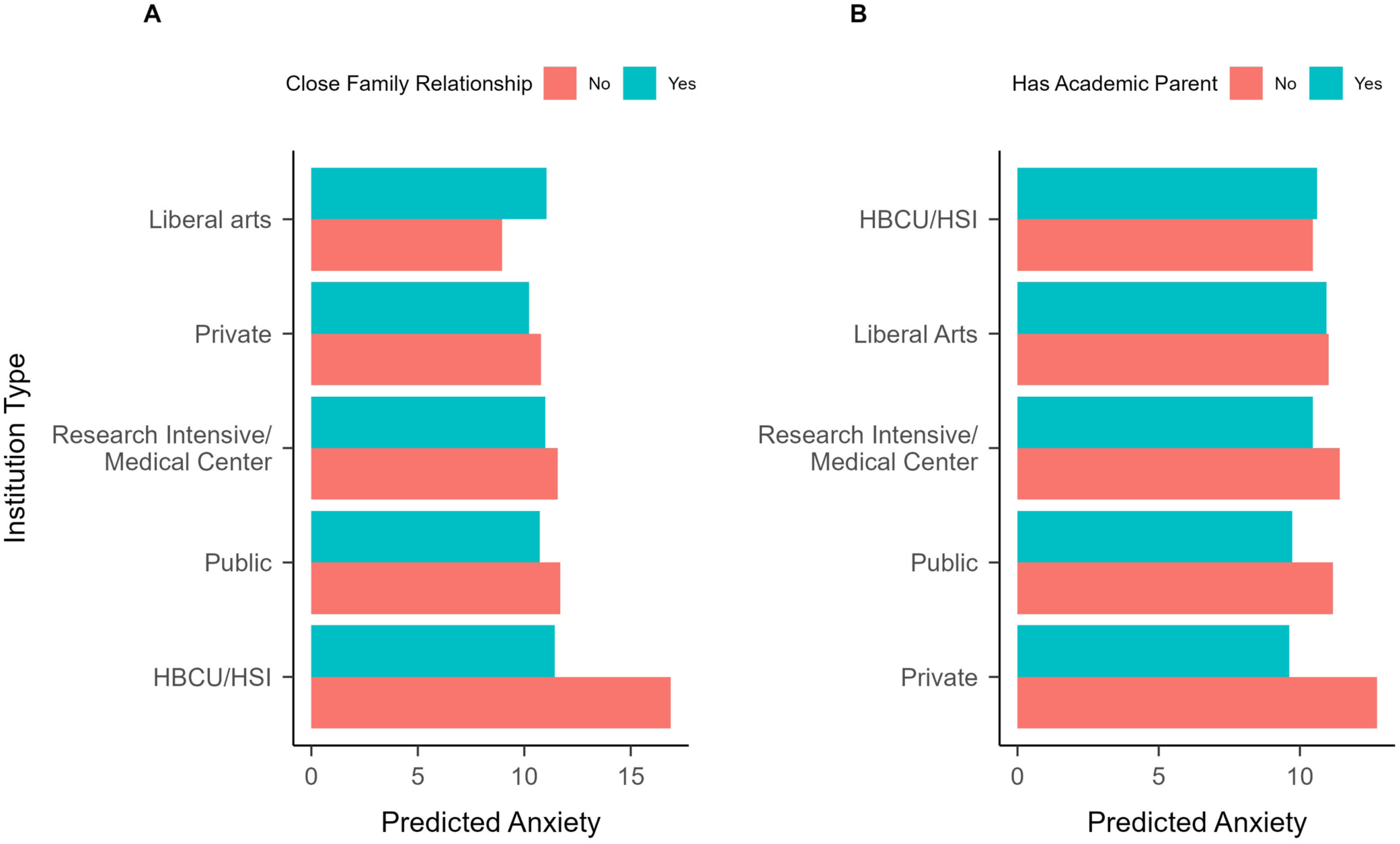
**Predicted Effect of Institution Type on Anxiety Based on Close Family Relationship or Academic Parent Status**. (A) Predicted anxiety levels based on close family relationships across different institution types. (B) Predicted anxiety levels based on having an academic parent across different institution types. Anxiety levels are measured by the GAD-7 and are predictions of the linear model. HBCU = Historically Black Colleges and Universities; HSI = Hispanic-Serving Institutions; Private = Private 4-year colleges and universities; Public = Public 4-year colleges and universities.

**Table 3.**
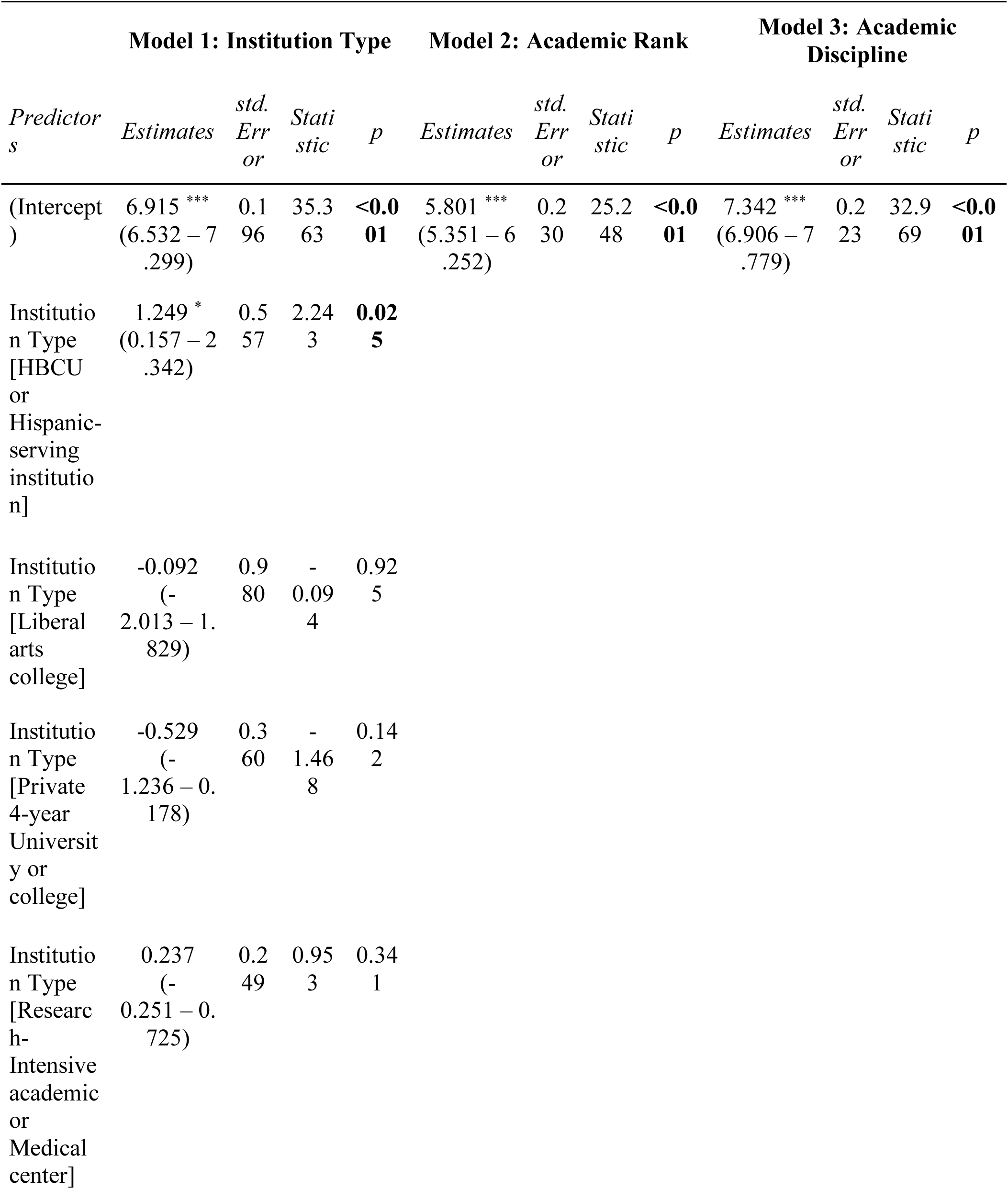
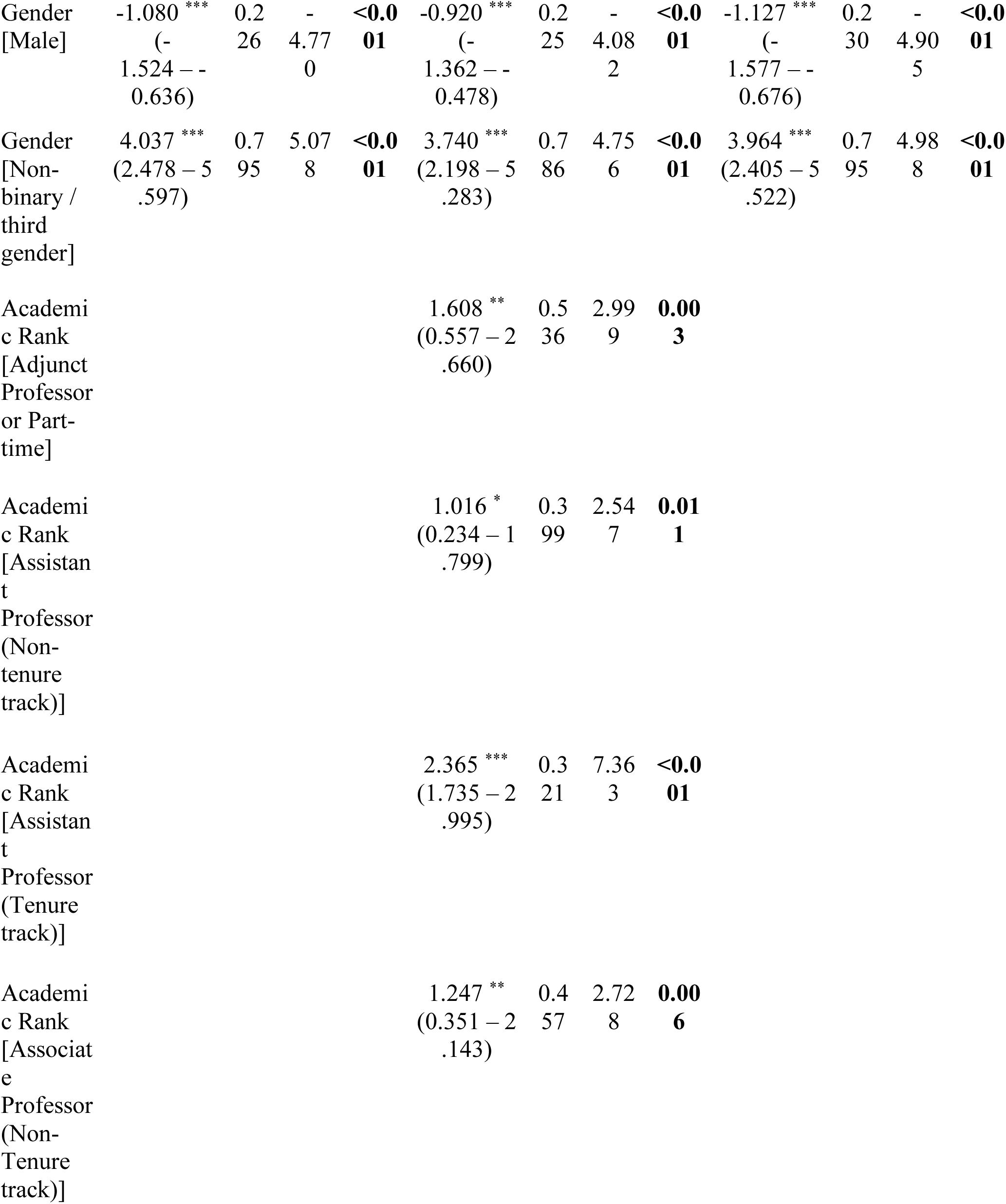
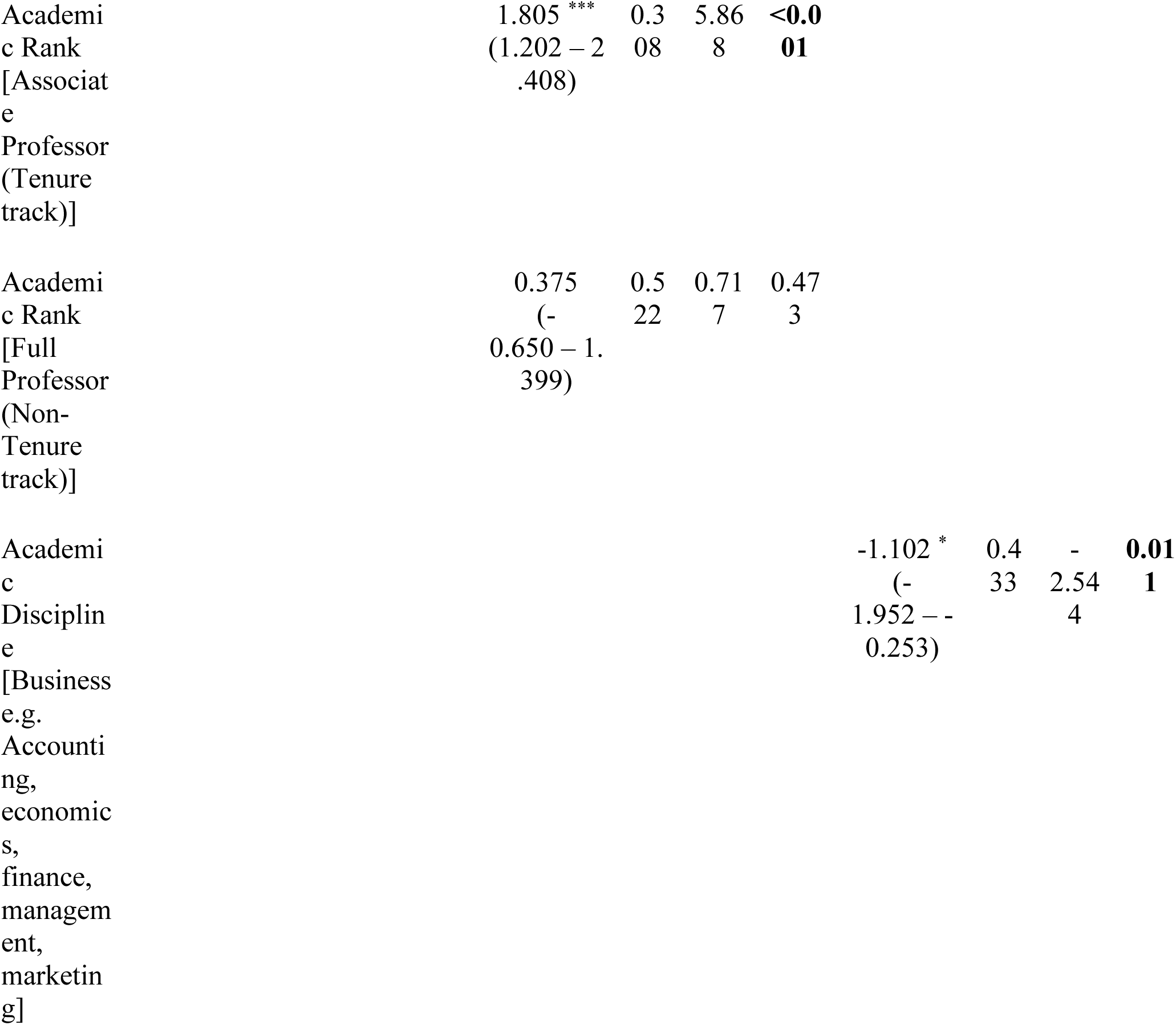
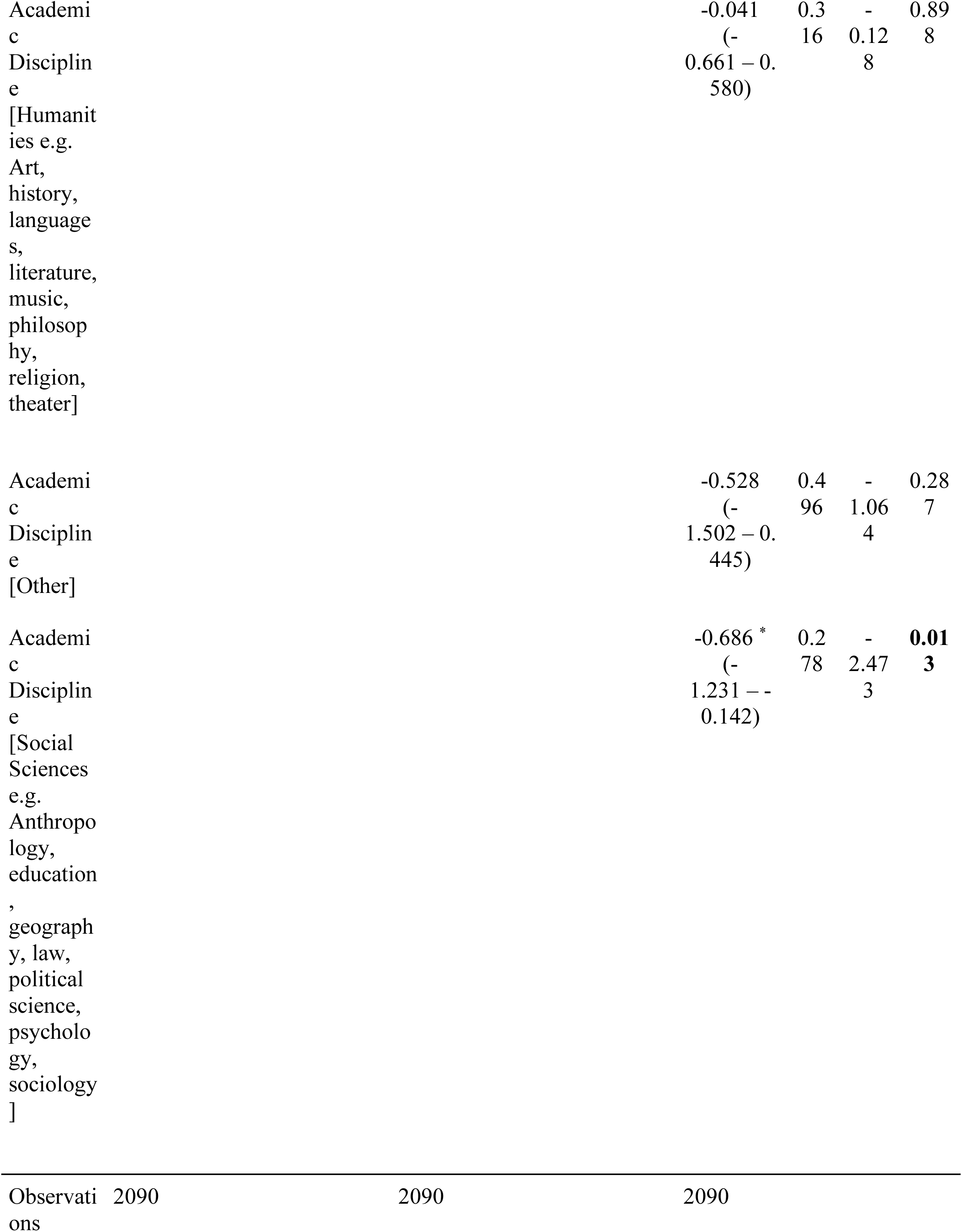
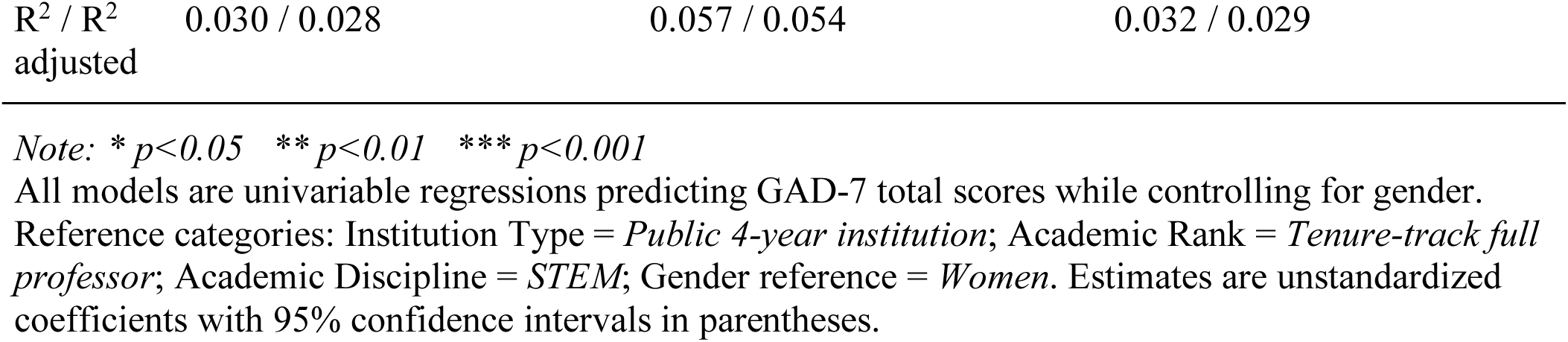
Univariable Regression Models Predicting GAD Scores, controlling for Gender.

### Does having a close family relationship or academic parent moderate the relationship between institution type and anxiety?

We examined close family relationships as a moderator of the relation between institution type and anxiety. The model was significant overall, F(11,2078) =7.55, p < .001, indicating that there were differences in the effect of close family relationships on anxiety across institution types, controlling for gender; however, effect sizes were small for institution type, η^2^ = .005, close family relationships, η^2^ = .004, and their interaction, η^2^ = .005. Relative to faculty at public institutions, there was a significant effect of close family relationships on anxiety for faculty at HBCU/HSIs, such that faculty at HBCUs/HSIs were significantly more anxious when they did not have close family relationships. For faculty at public institutions (reference group), there was also a significant main effect of close family relationships easing anxiety. When family relationships were close, HBCU/HSI faculty were less anxious did not have a significant interaction with any institution types, however, ps > .272.

The model for the effect of academic parent as a moderator of the relationship between institution type and anxiety was significant overall, F(11,2078) =6.75, p < .001. Similar to the previous model, effect sizes for main effects and interactions were small: institution type, η^2^ = .005, academic parent, η^2^ = .002, and their interaction, η^2^ = .003. No interactions were significant, suggesting that having or not having a parent who is an academic does not result in a significant change in anxiety at different institution types and that significance of the overall model is likely driven by the gender effect on anxiety. All effects can be viewed in Table 4.

**Table 4.**
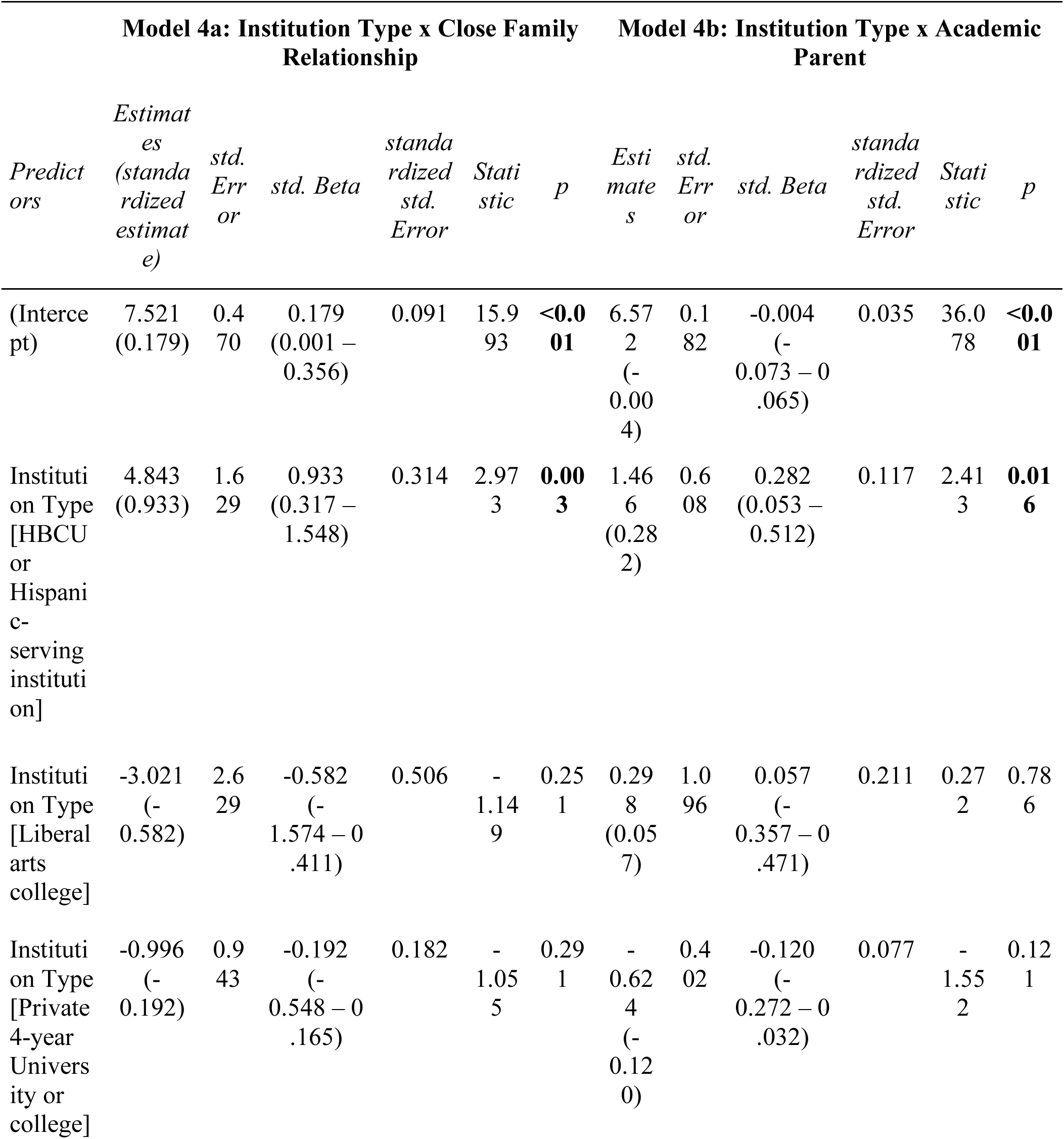
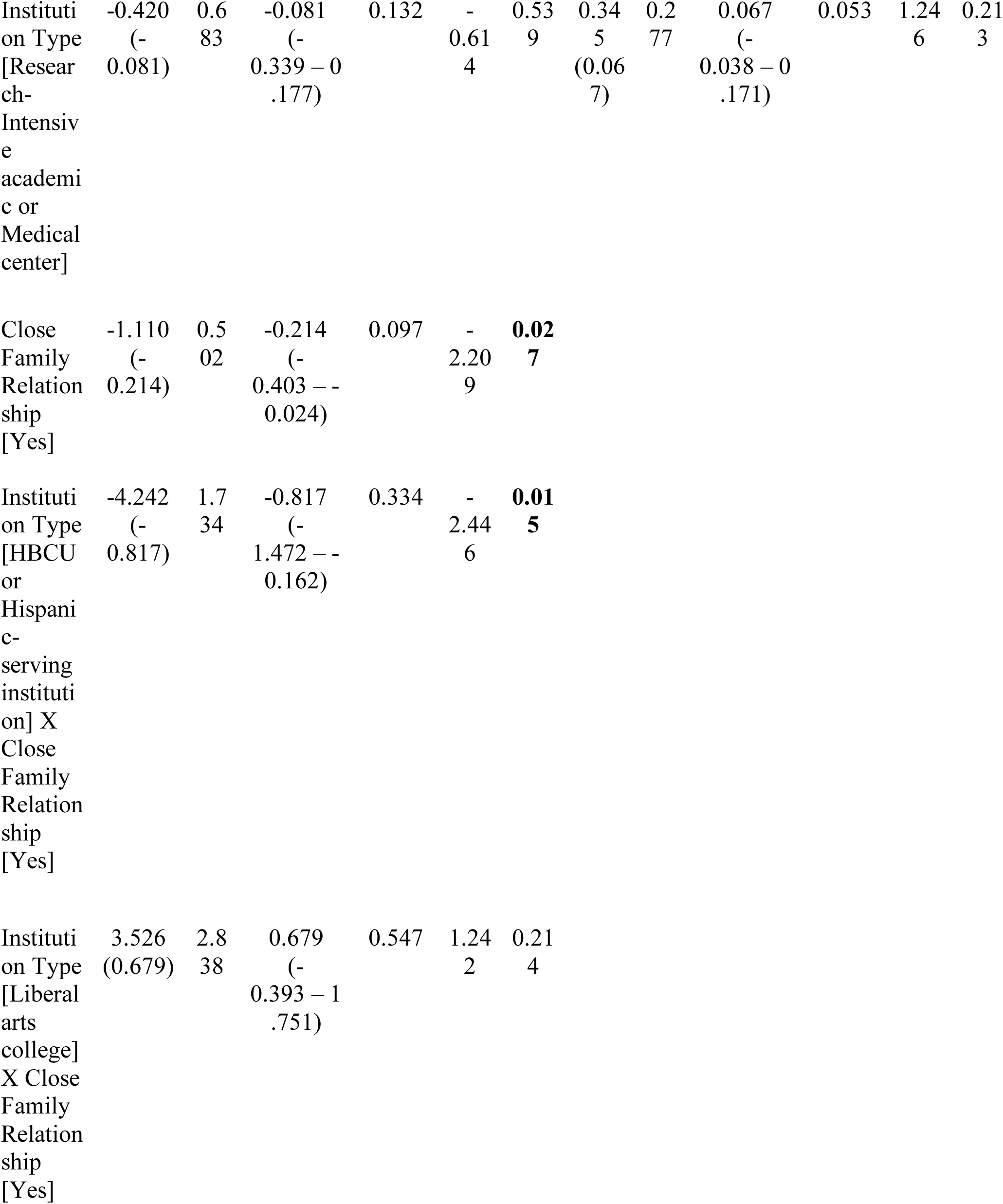
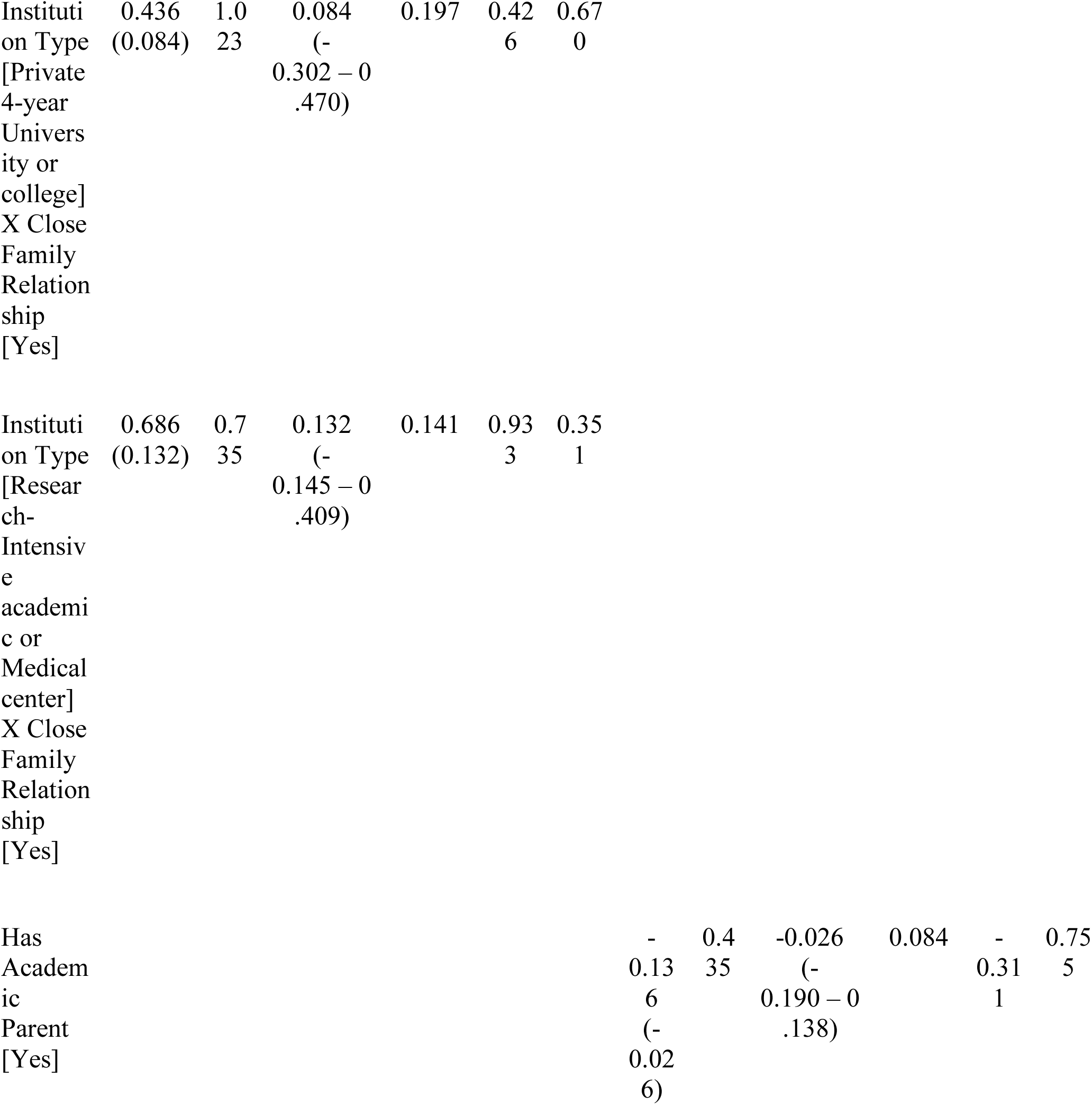
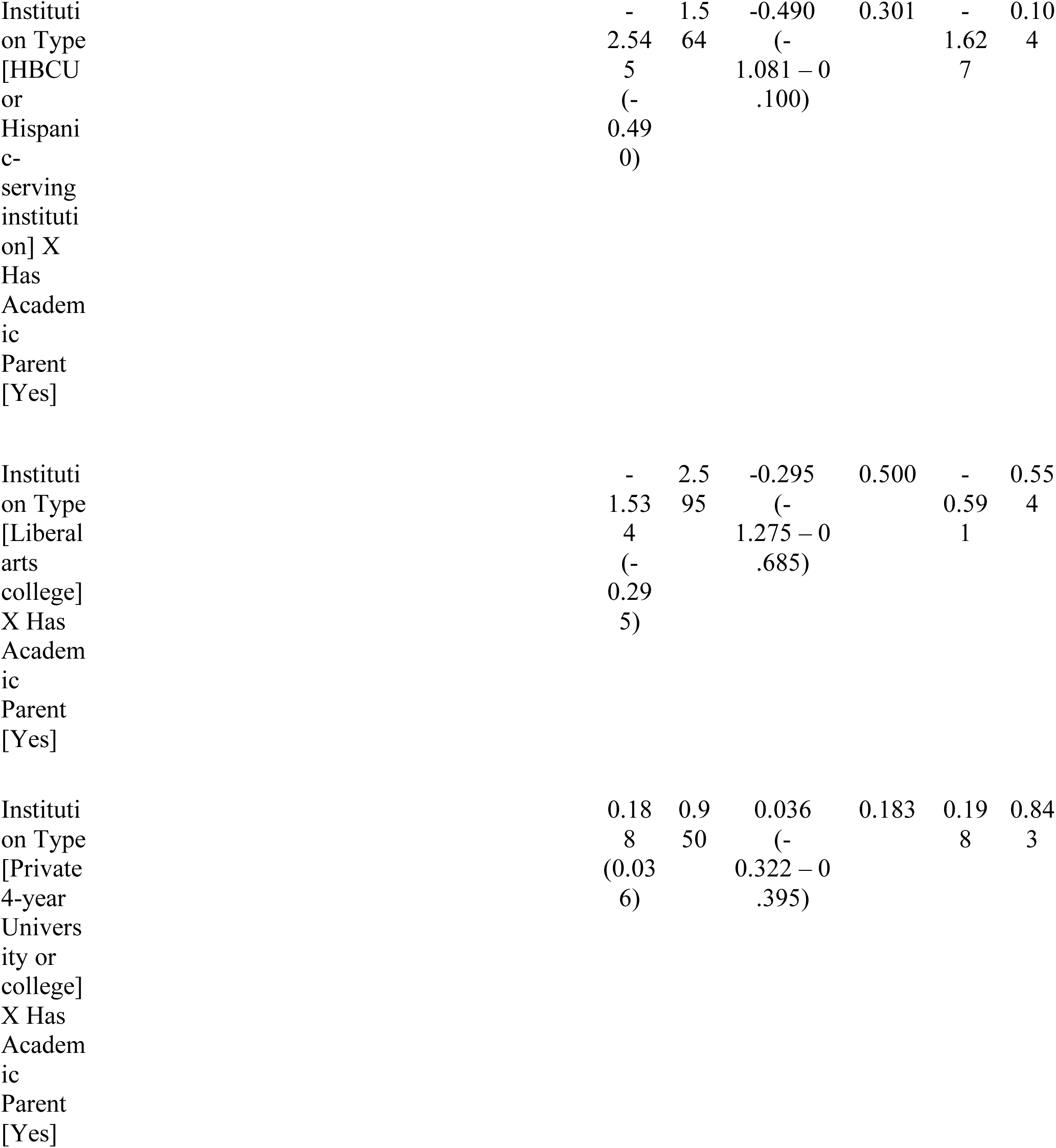
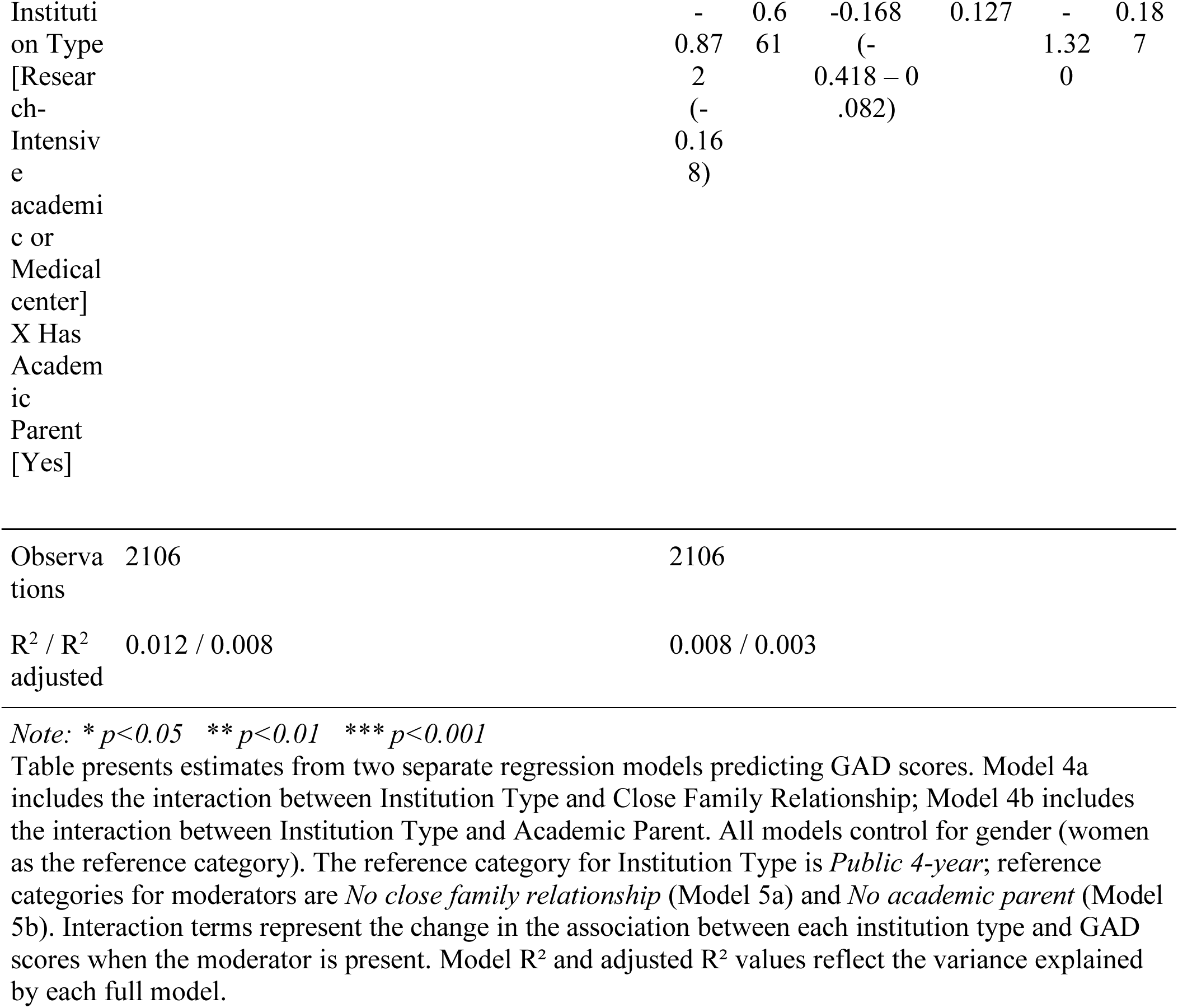
Regression Models Predicting GAD Scores for Institution Type.

### Does having close family relationships or an academic parent moderate the relationship between academic rank and anxiety?

The model for close family relationships’ moderation of the effect of academic rank on anxiety was significant overall, F(15,2074) =9.51, p < .001, indicating that there were differences in the effect of close family relationships on anxiety across academic ranks, controlling for gender; however, effect sizes were small for academic rank, η^2^ = .04, close family relationships, η^2^ = .003, and their interaction, η^2^ = .005. Close family relationships significantly lowered anxiety among adjunct/part-time professors and among non-tenure track full professors relative to tenure-track full professors (Figure 2). The model for having an academic parents’ moderation of the effect of academic rank on anxiety was also significant overall, F(15,2074) = 9.25, *p < .*001 with small effect sizes for main effects of academic rank, η^2^ = .04, academic parent, η^2^ = .001, and their interaction, η^2^ = .005. There was a significant effect of having an academic parent on lowering anxiety among adjunct professors. No other interactions were significant. All effects can be viewed in Table 5.

**S Figure 2.**
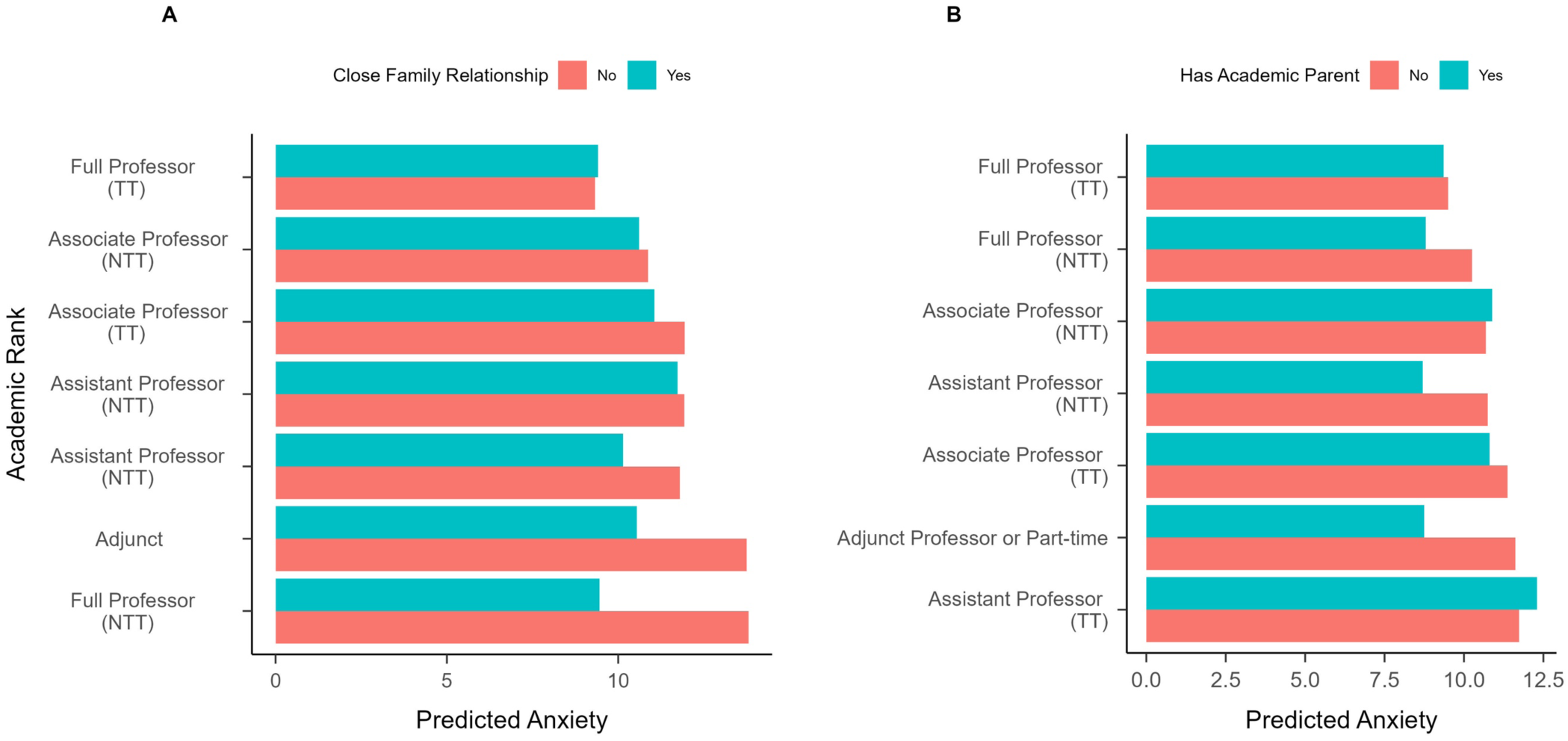
Predicted Effect of Academic Rank and Tenure on Anxiety Based on Close Family Relationship or Academic Parent Status. (A) Predicted anxiety levels based on close family relationships across different academic ranks. (B) Predicted anxiety levels based on having an academic parent across different academic ranks. Anxiety levels are measured by the GAD-7. TT = Tenure Track; NTT = Non-tenure Track. Adjunct represents adjunct and part-time faculty.

**Table 5.**
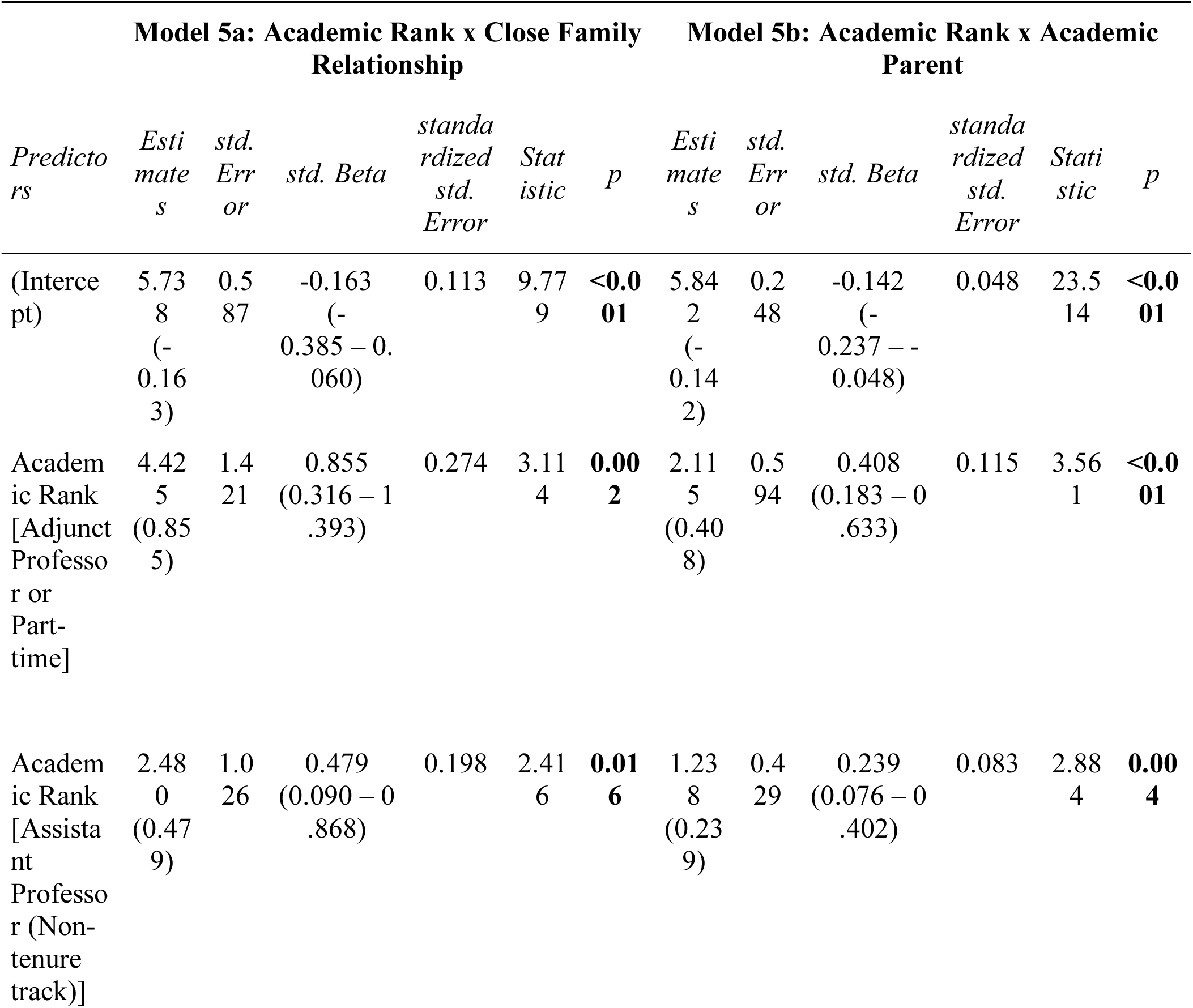
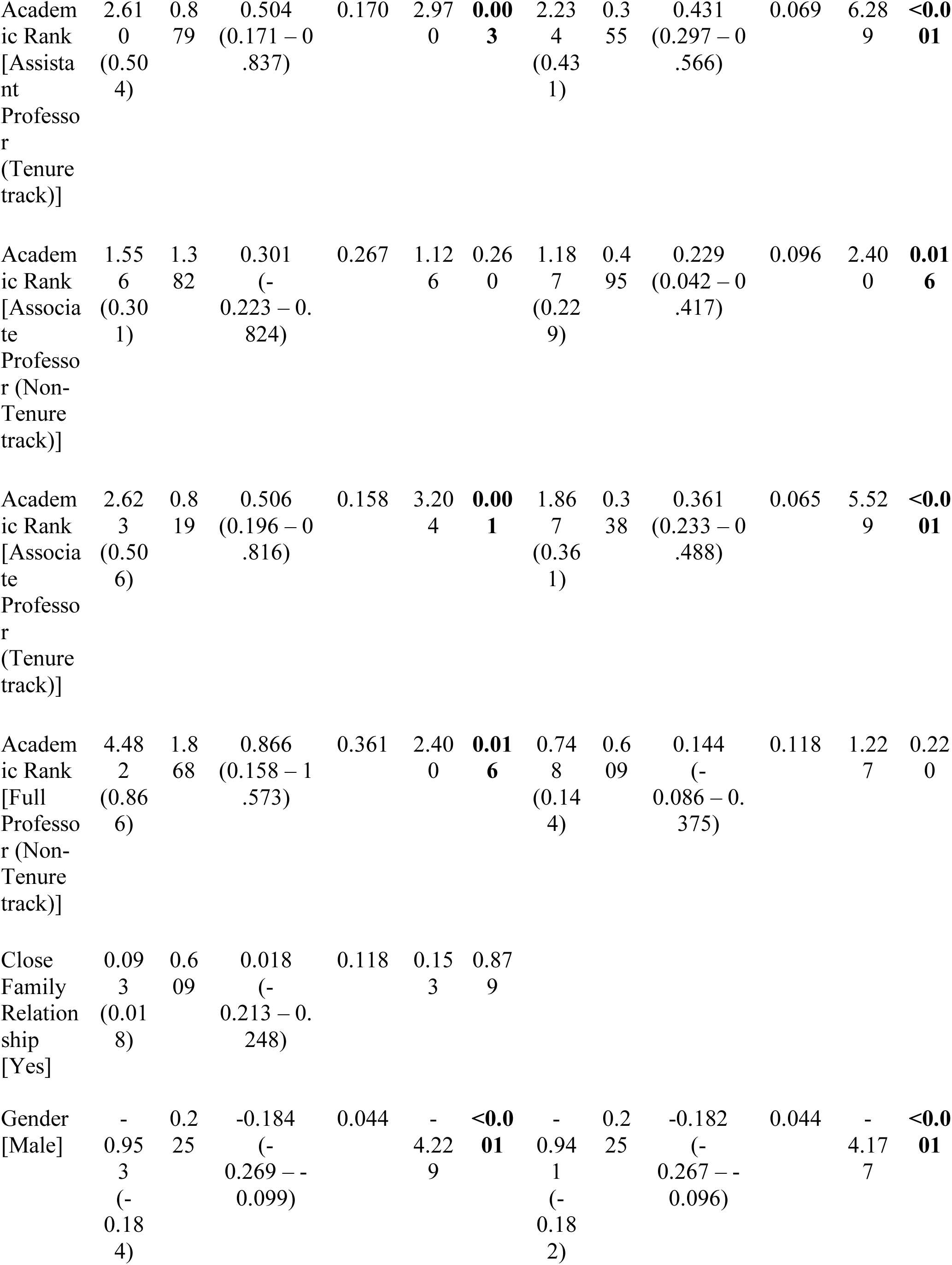
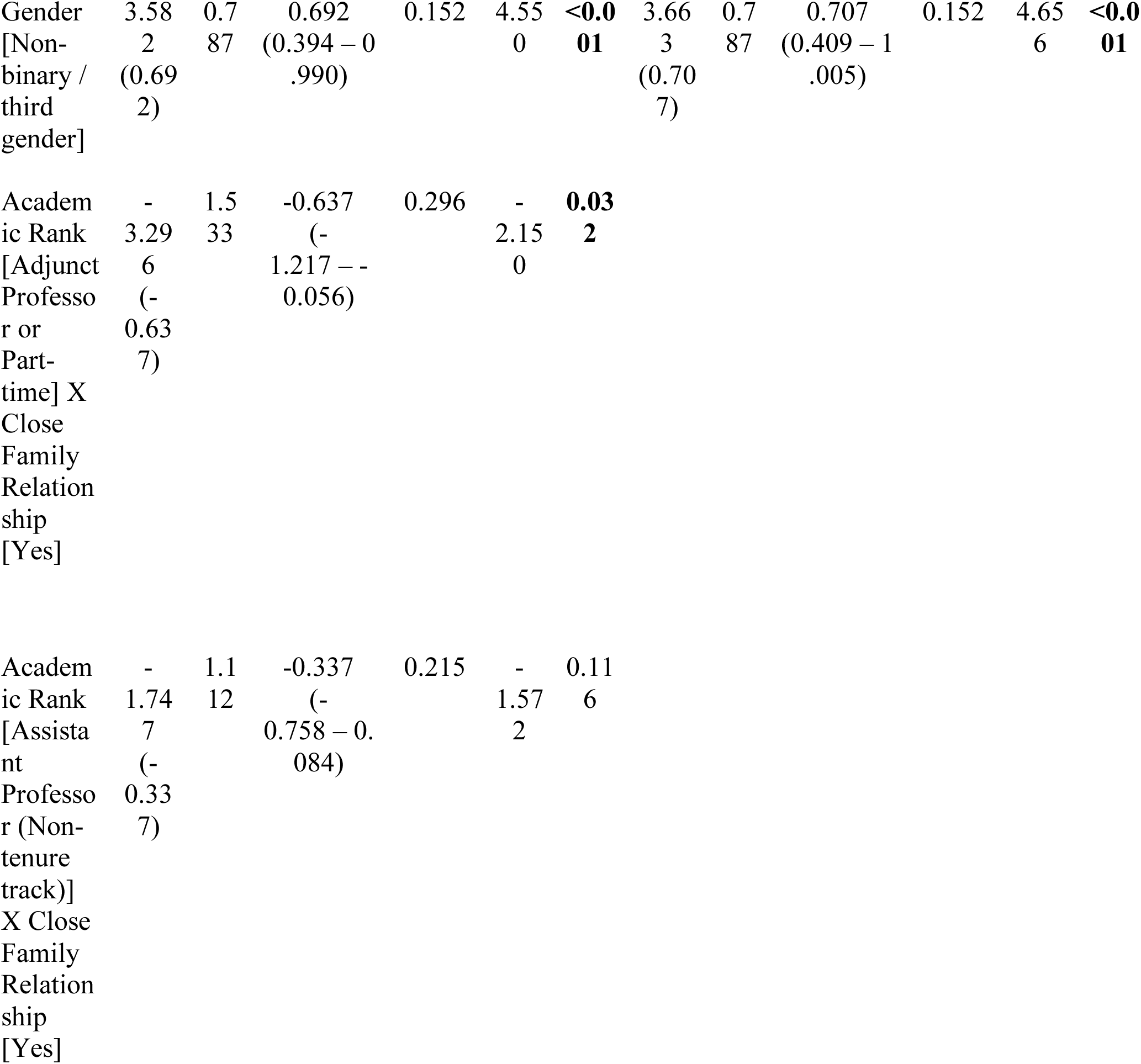
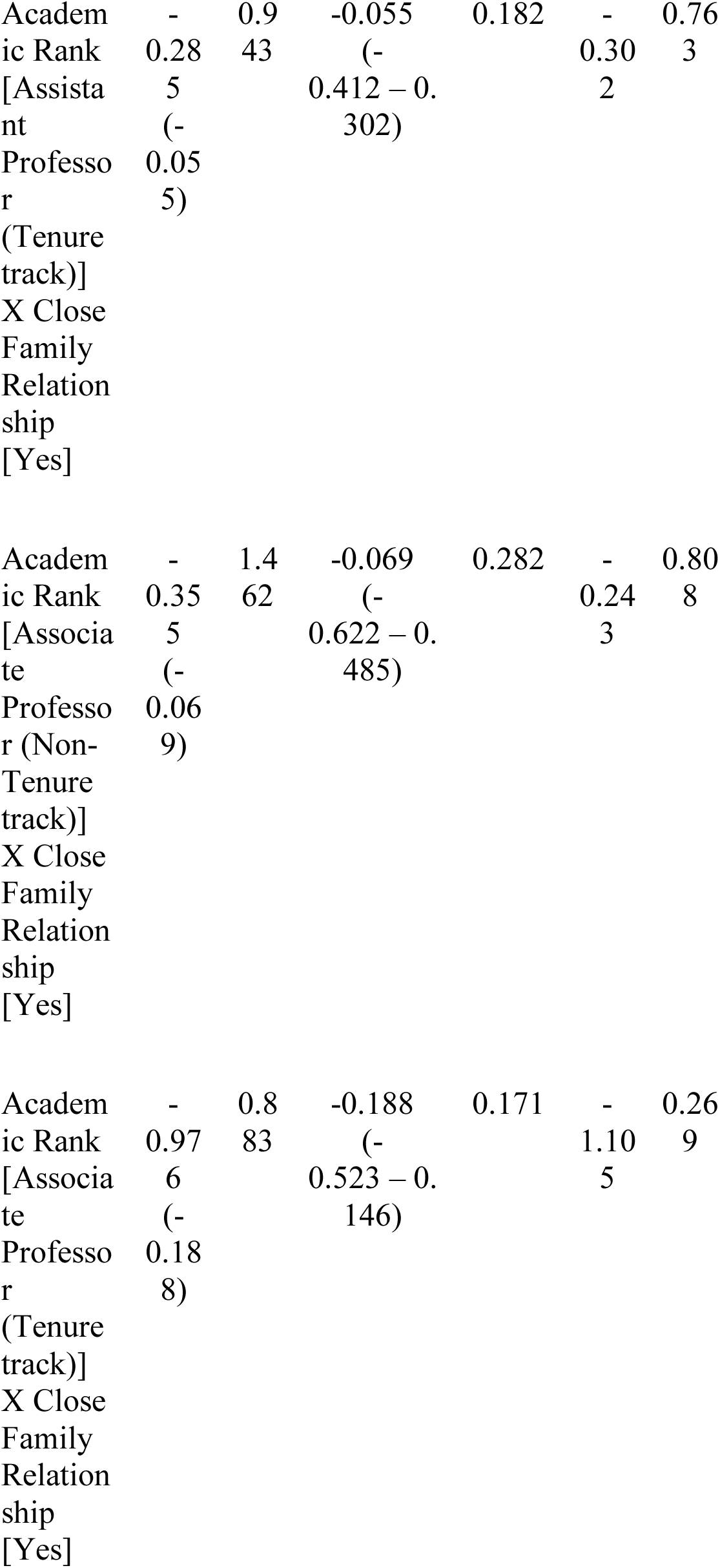
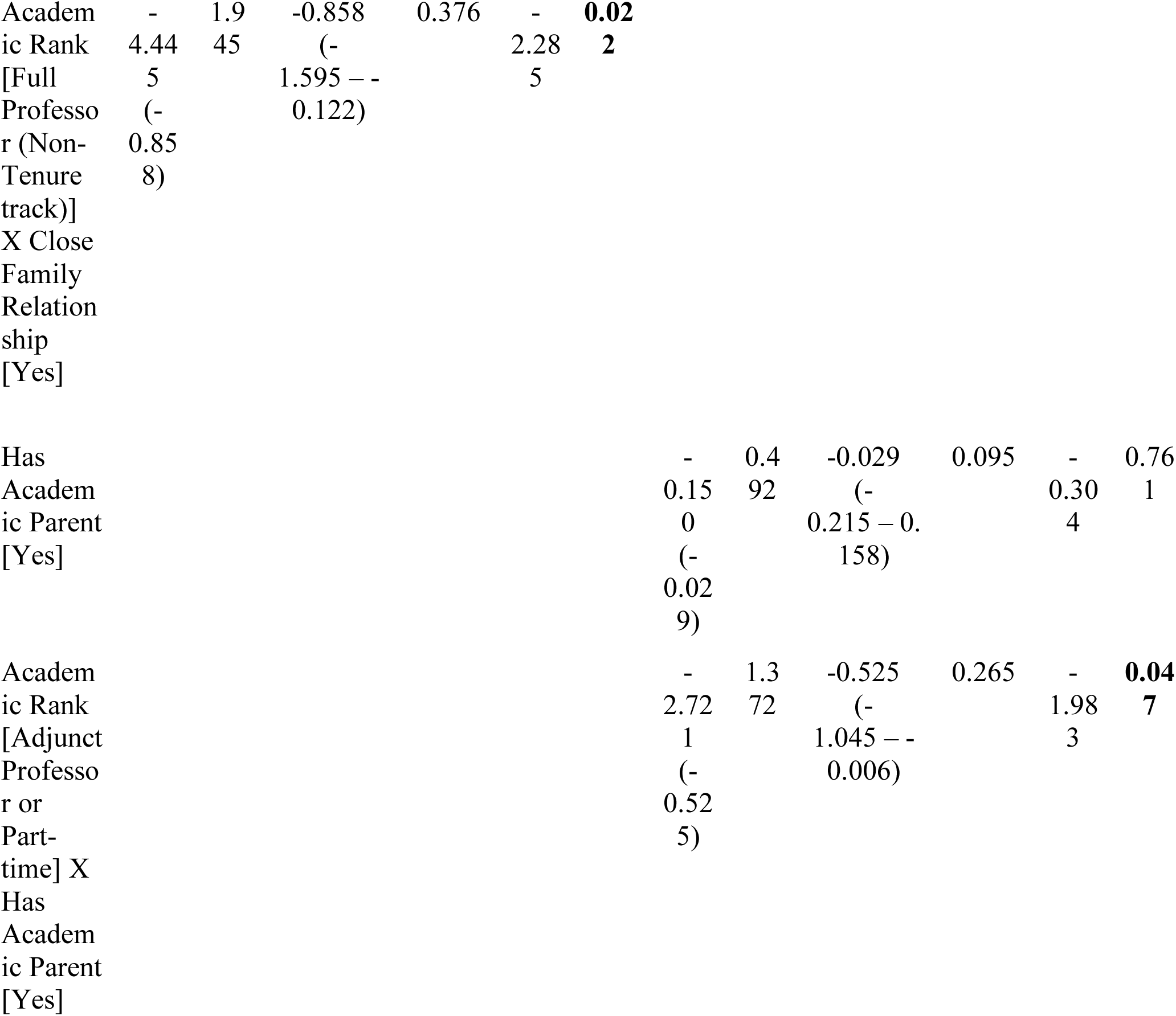
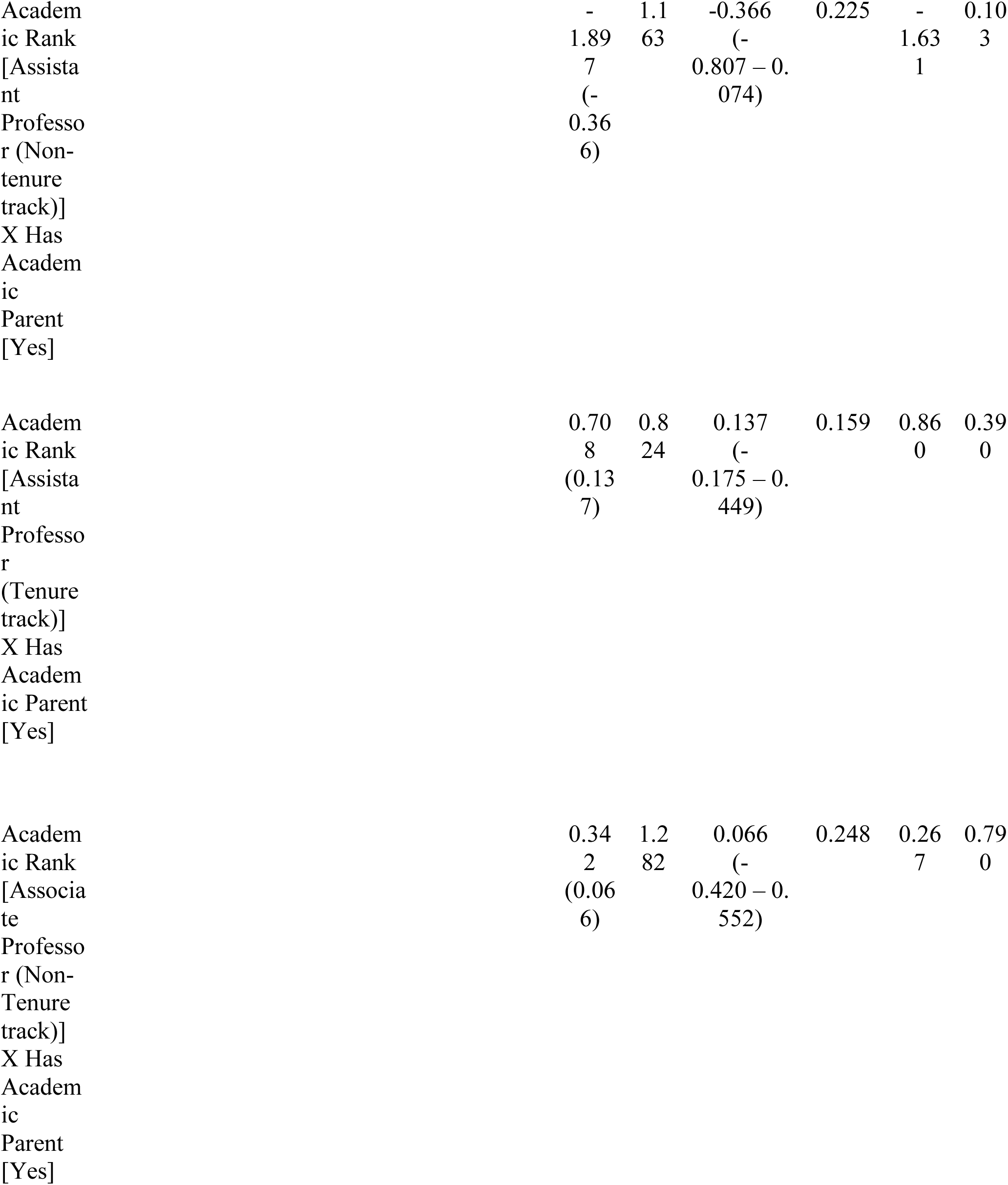
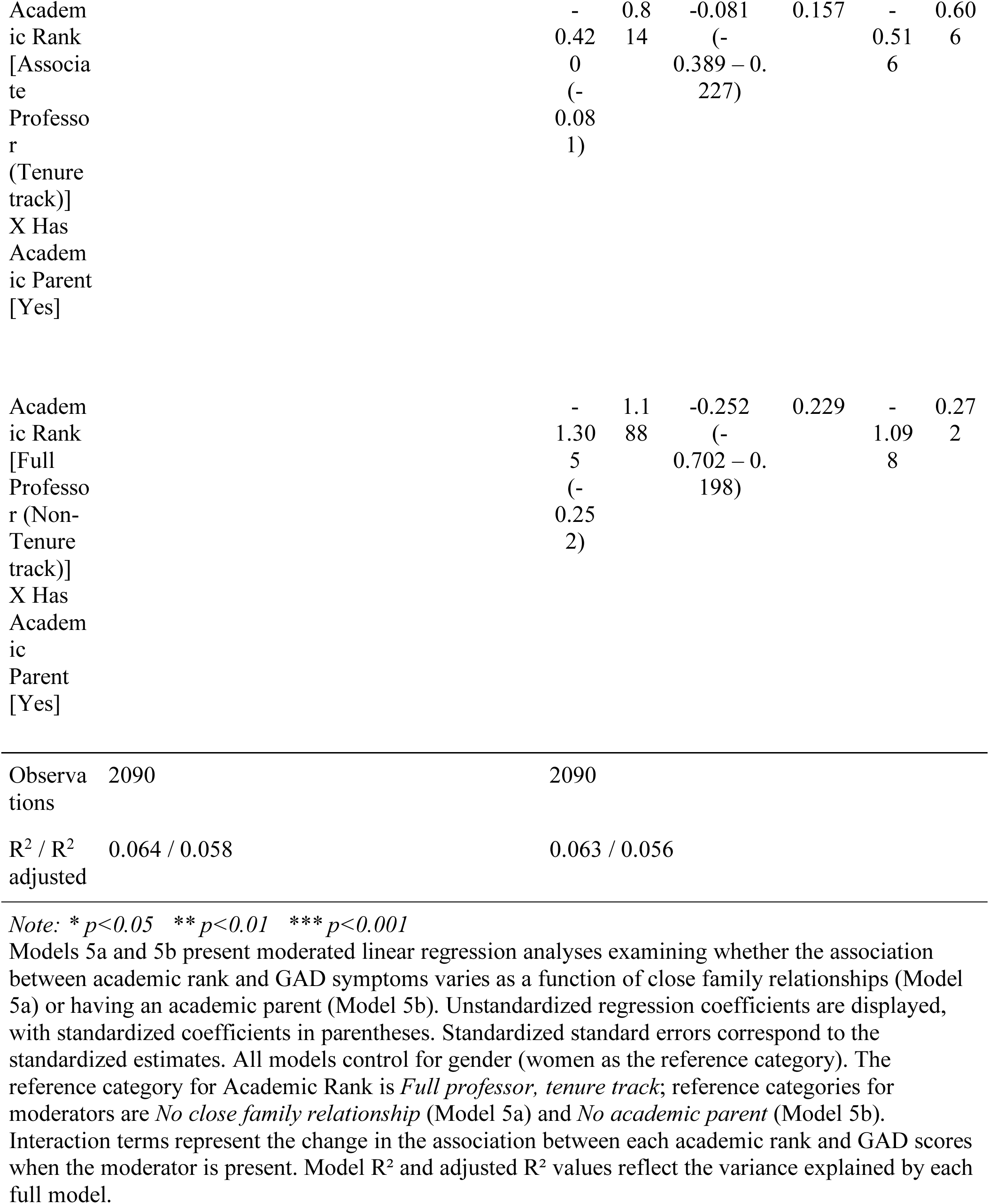
Regression Models Predicting GAD Scores with moderated Academic Rank.

### Does having close family relationships or an academic parent moderate the relationship between academic discipline and anxiety?

Lastly, we examined the effect of familial relationships on anxiety by academic discipline (Figure 3). The model for close family relationships’ moderation of the effect of academic rank on anxiety was significant overall, F(15, 2074) =9.51, p < .001, indicating that there were differences in the effect of close family relationships on anxiety across academic ranks, controlling for gender; however, effect sizes were small for academic discipline, η^2^ = .006, close family relationships, η^2^ = .004, and their interaction, η^2^ = .0003. Close family relationships did not moderate the effect of being a member of any specific academic disciplines on anxiety relative to STEM faculty. The model for academic parents’ moderation of the effect of academic discipline on anxiety was significant overall, F(11, 2078) = 6.6, p < .001. Similar to the previous model, effect sizes were small for academic discipline, η^2^ = .005, academic parent, η^2^ = .002, and their interaction, η^2^ = .0008. Across disciplines, having an academic parent did not have a significant impact on anxiety. All effects can be viewed in Table 6.

**S Figure 3.**
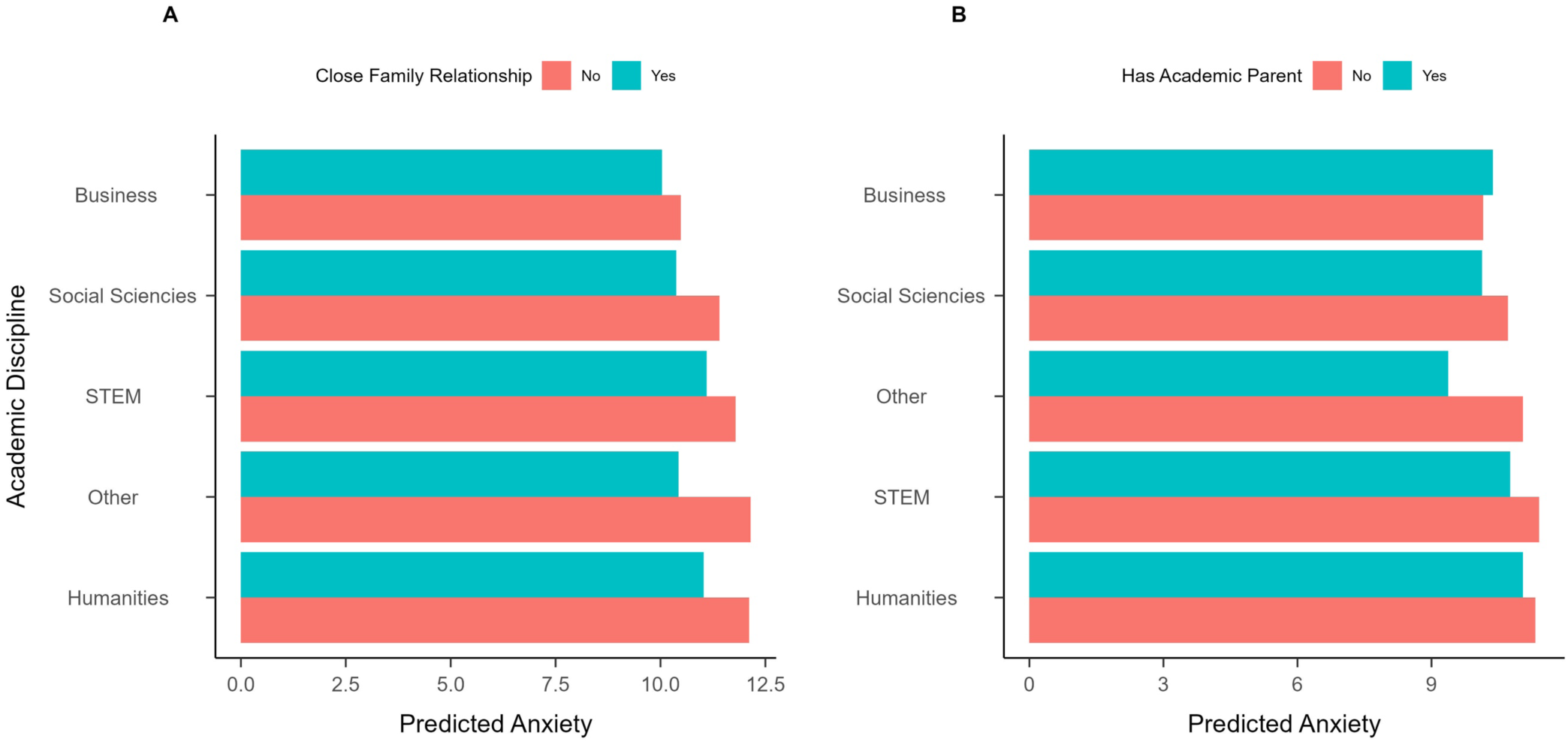
**Predicted Effect of Academic Discipline on Anxiety Based on Close Family Relationship or Academic Parent Status**. (A) Predicted anxiety levels based on close family relationships across surveyed academic disciplines (B) Predicted anxiety levels based on having an academic parent across surveyed academic disciplines. Anxiety levels are measured by the GAD-7.

**Table 6.**
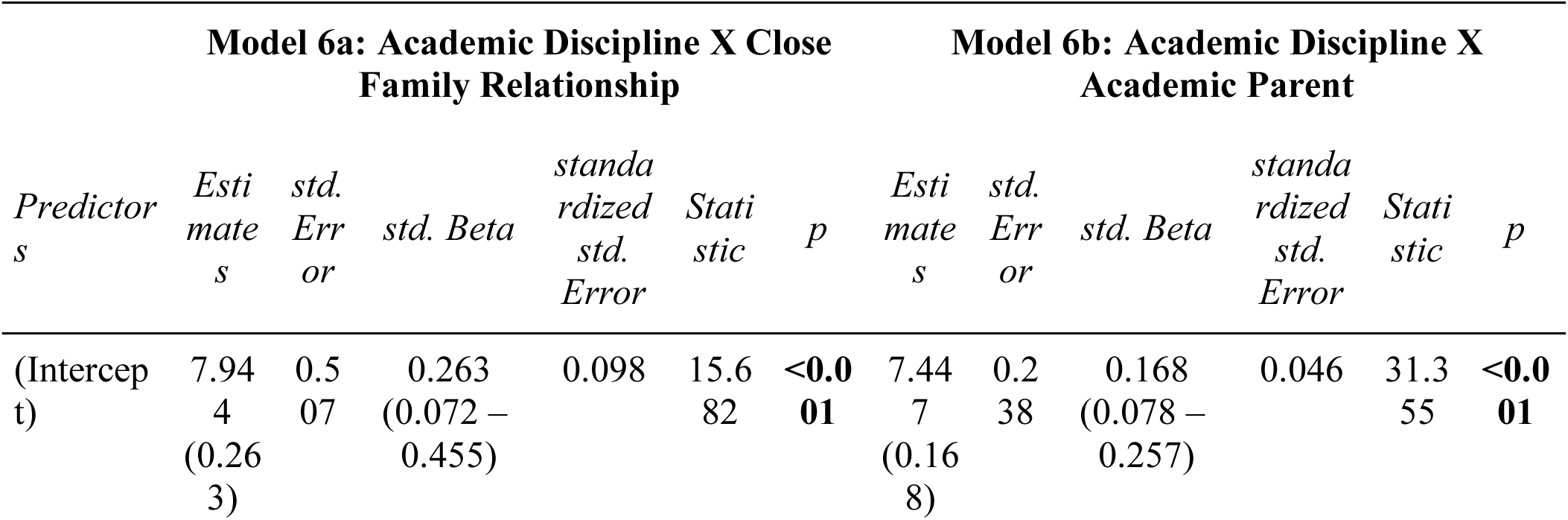
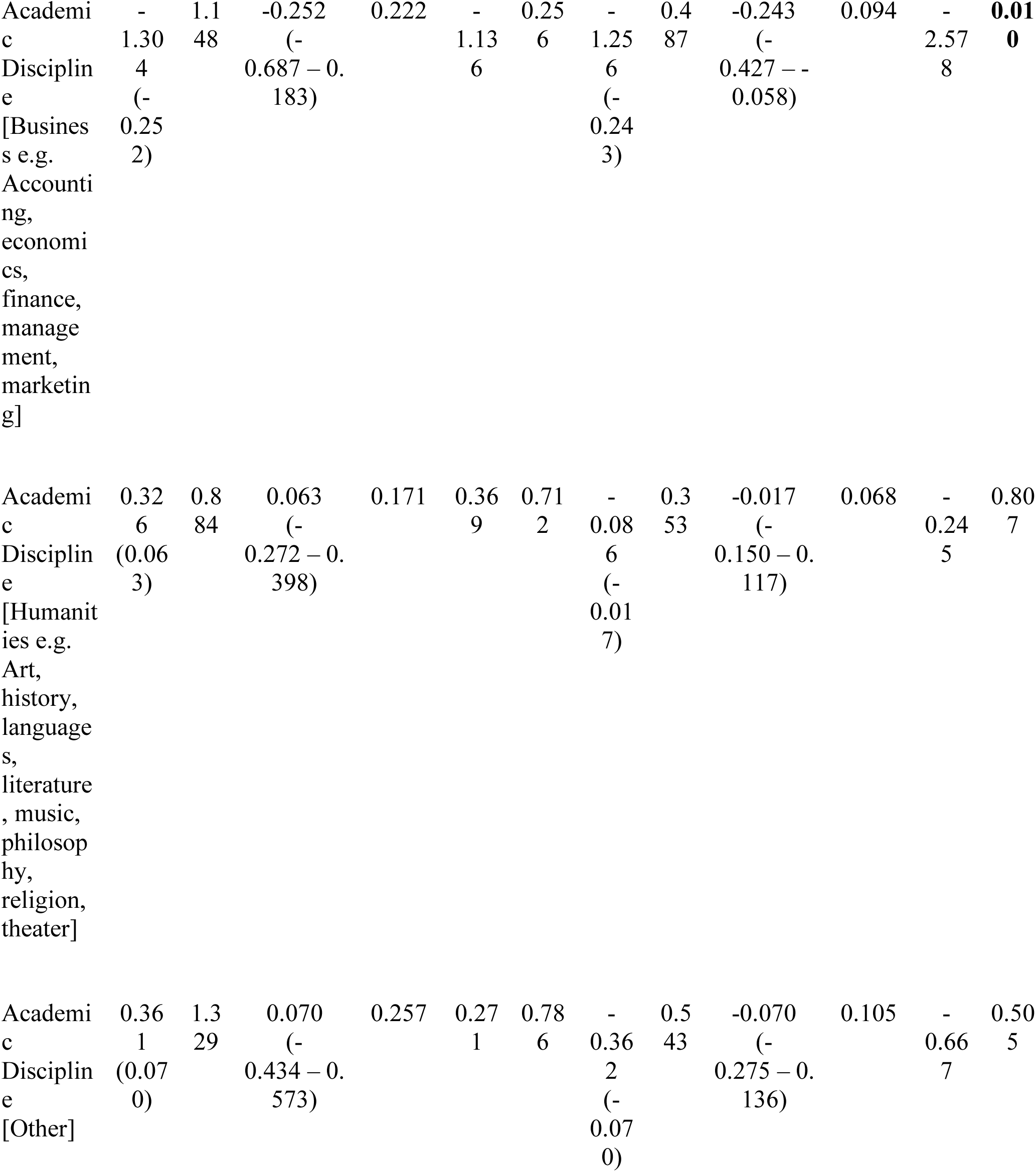
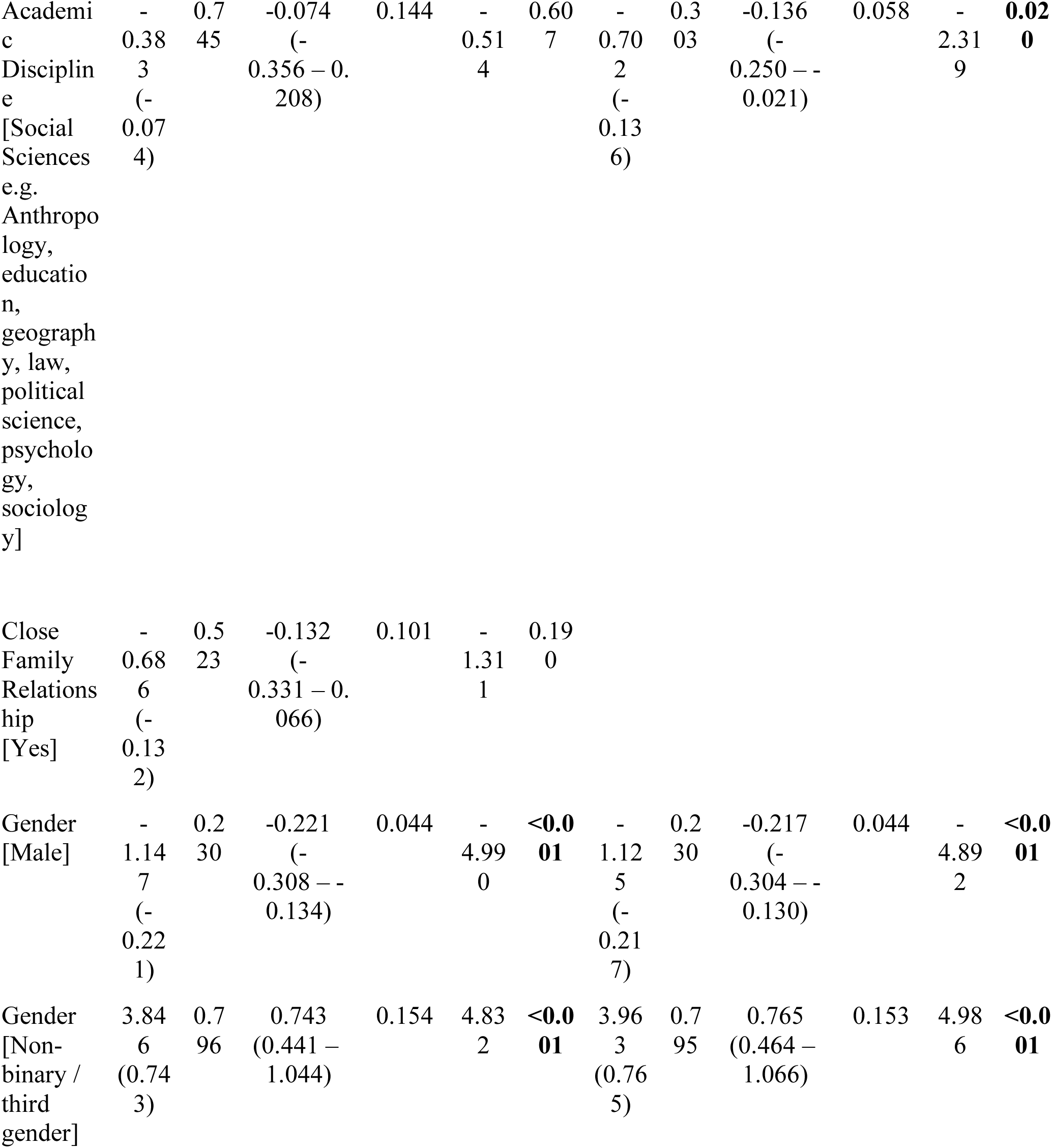
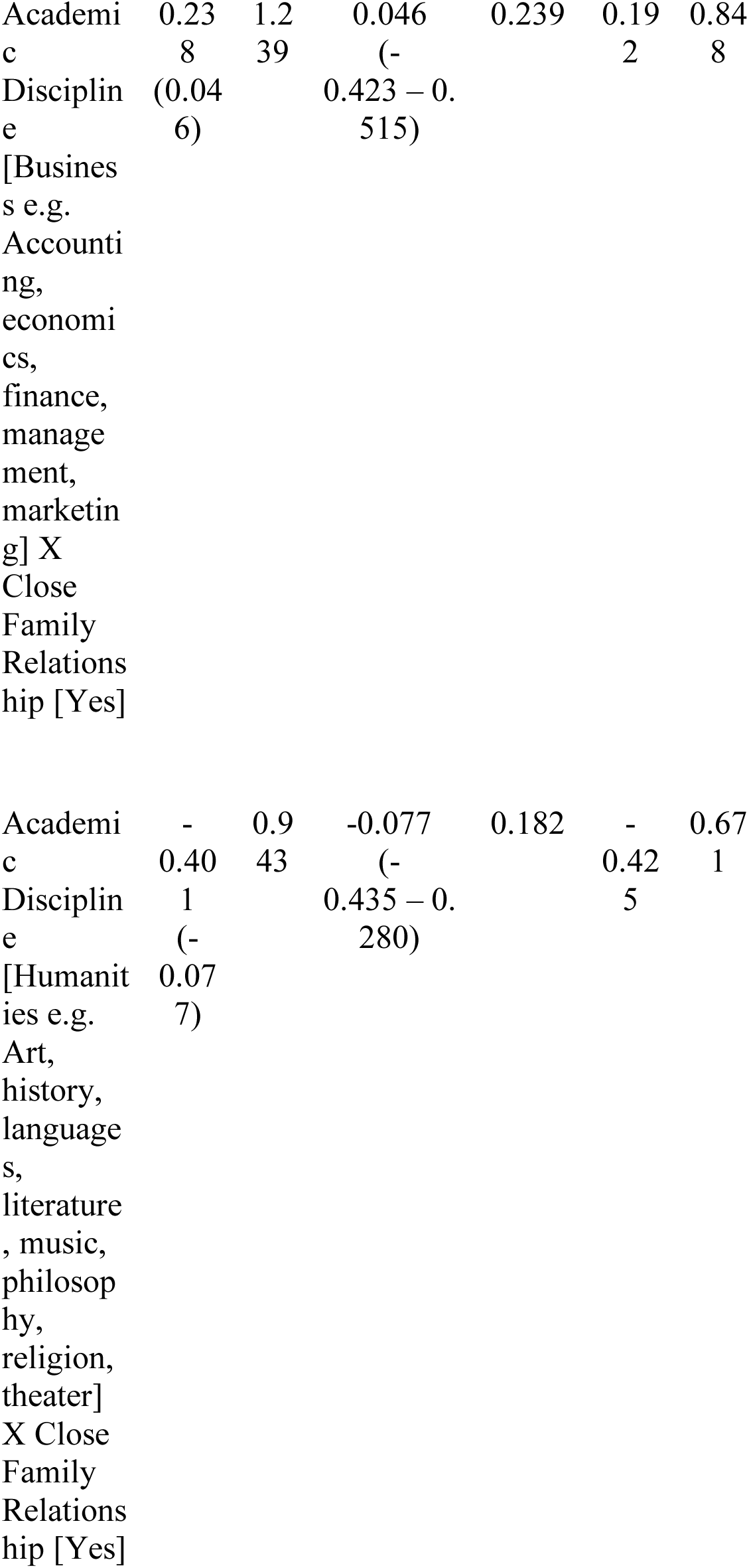
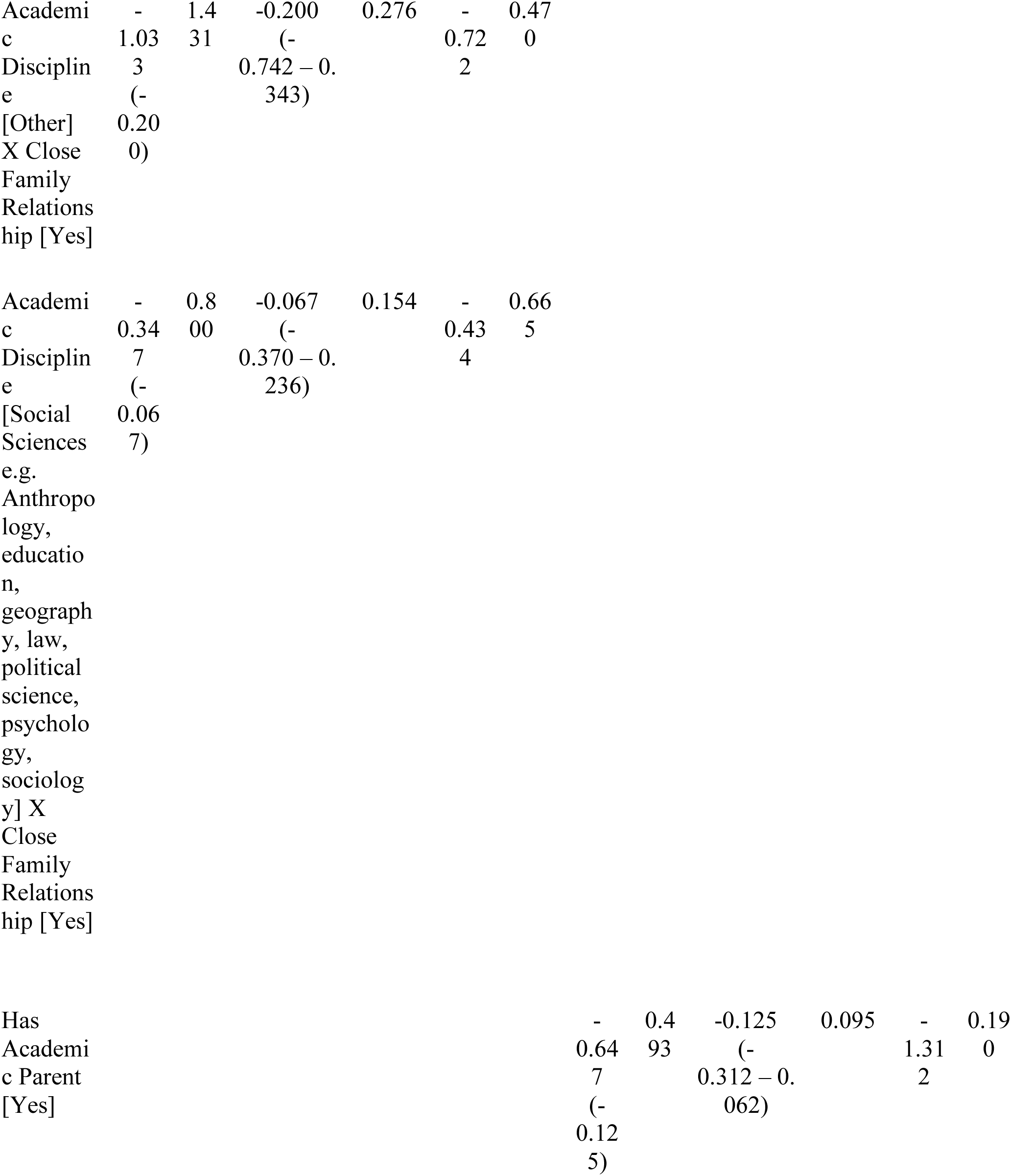
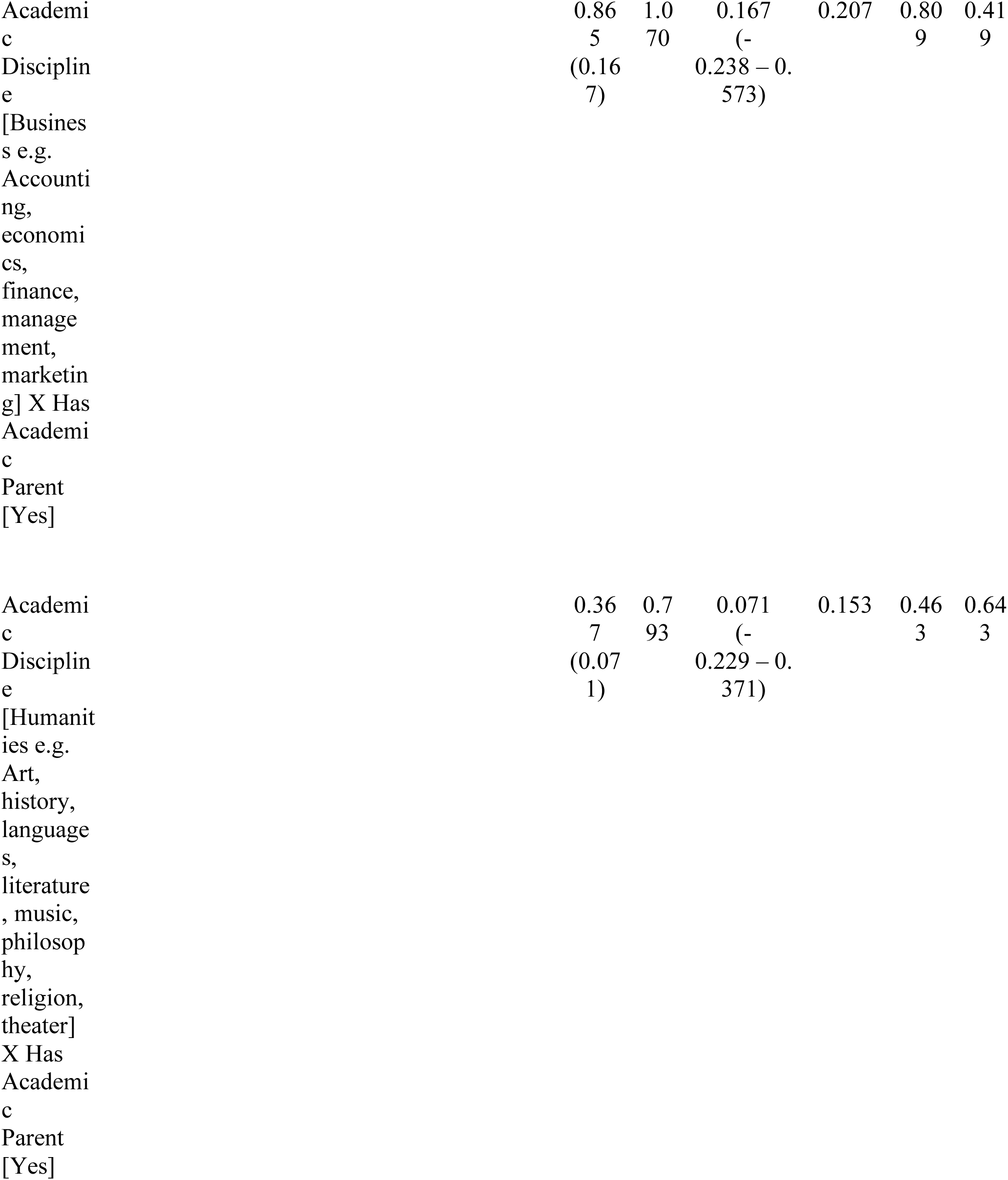
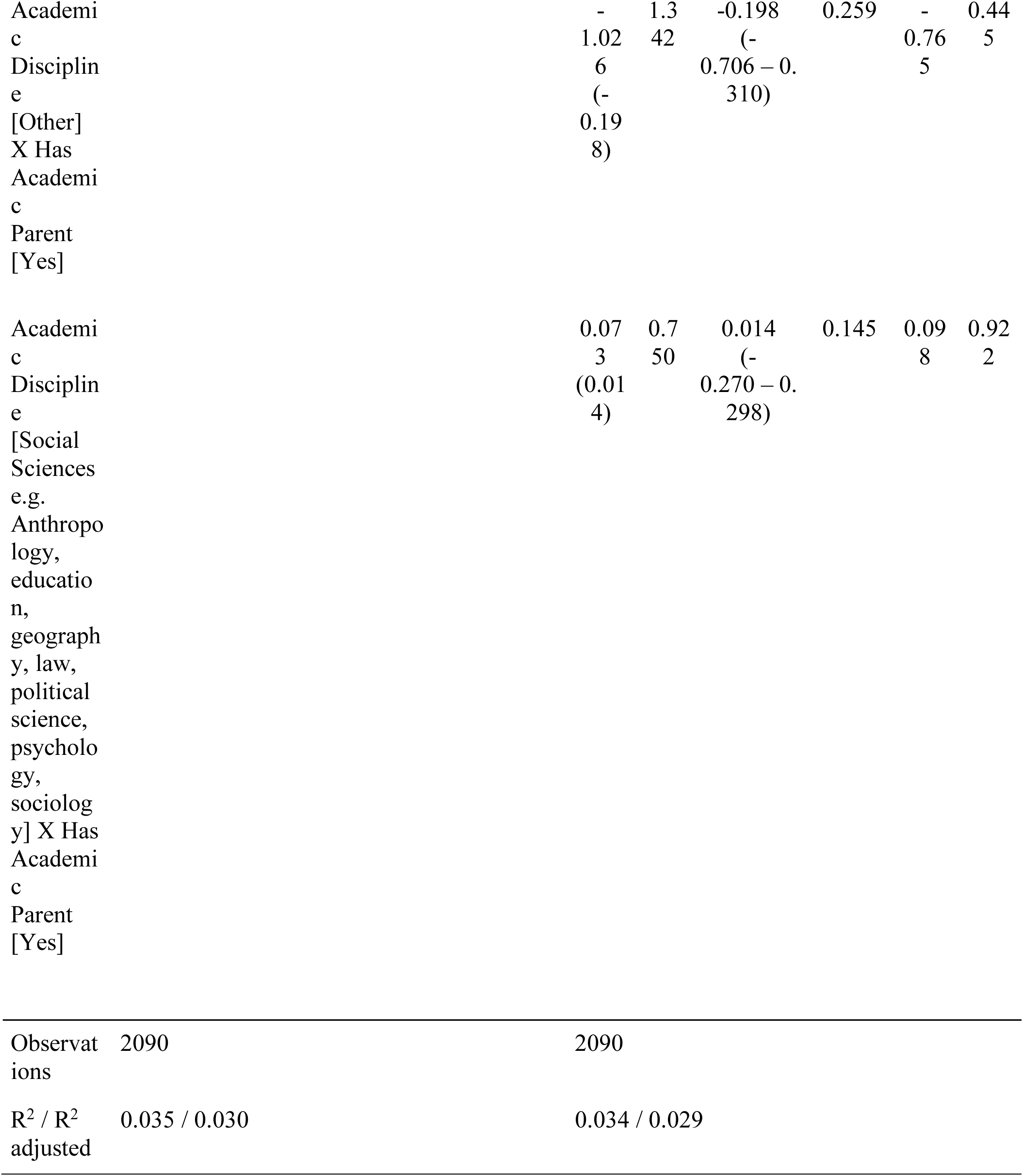
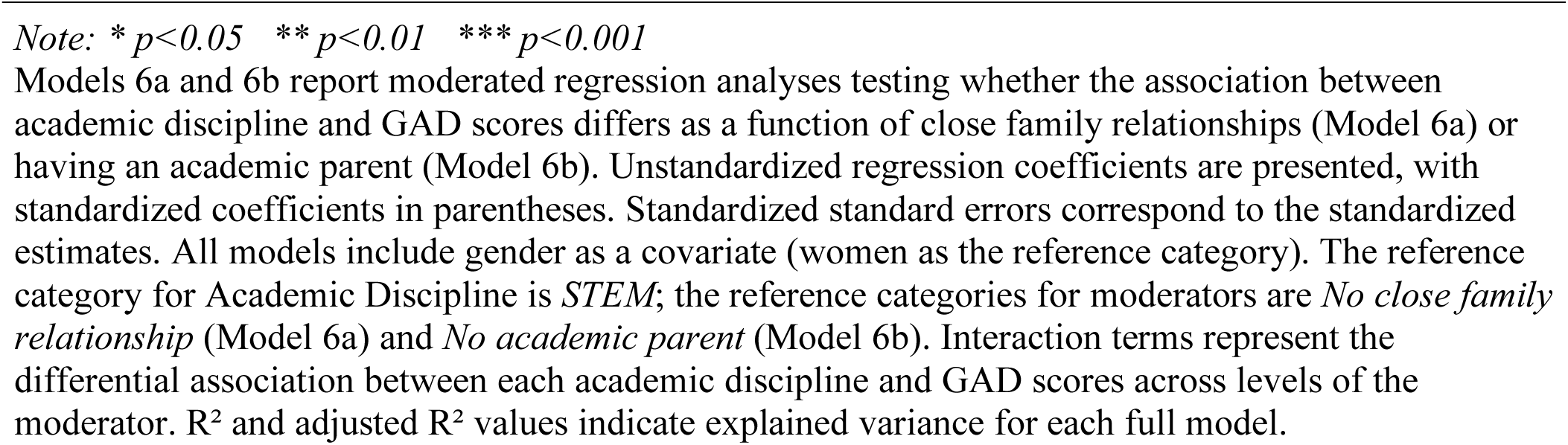
Regression Models Predicting GAD Scores with Moderated Academic Discipline.

## Discussion

Our study provides a large-scale examination of anxiety among U.S. academic faculty, revealing critical insights into how familial relationships and academic lineage moderate anxiety across different institutional contexts. These findings address a significant gap in our understanding of faculty mental health and offer actionable insights for institutional interventions.

The protective effect of close family relationships on faculty anxiety, particularly strong at HBCUs/HSIs, suggests that social support systems play a crucial role in faculty mental well-being. This finding extends previous research on social support in academic settings^21^ by demonstrating its specific impact on faculty anxiety levels. The pronounced effect at HBCUs/HSIs may reflect these institutions’ historically strong emphasis on community and mentorship^22,23^, suggesting that institutional culture can amplify the benefits of family support. Our finding that having an academic parent has impacts for adjunct faculty as a protective factor reveals a previously unexamined dimension of academic mental health. This finding contextualizes research showing that tenure-track faculty are 25 times more likely to have a parent with a PhD^24^, suggesting that having an academic parent may confer advantages to degree-seeking behavior, matriculation, or securing an academic job; however, these effects may be primarily felt by adjunct/part-time faculty. The clear relationship between academic rank and anxiety levels, with tenure-track assistant professors showing the highest anxiety scores, highlights a critical vulnerability in academic career trajectories. This finding confirms that the traditional tenure system, while designed to ensure academic excellence^25^, creates a significant psychological burden for early-career faculty. The relatively lower anxiety levels among non-tenure track full professors compared to their tenure-track counterparts raises important questions about the relationship between job security and mental well-being in academia^12^.

The minimal impacts of academic discipline on anxiety challenges assumptions about discipline-specific stress levels in academia. Instead, our findings suggest that institutional factors and support systems play small, but consistent roles in faculty mental health. This insight argues for institutional interventions that focus on building supportive environments rather than discipline-specific solutions. It also suggests further research into other types of social support as buffers for anxiety among faculty. For example, it may be that other details of familial relationships – whether parents were authoritative or permissive or whether close family relationships also included financial or emotional supports – may have lead to strong effects on anxiety and warrant future investigation.

Several limitations should be considered when interpreting these results. First, our study’s cross-sectional design prevents causal inference about the relationship between family support and anxiety levels. Second, the self-reported nature of the GAD-7 assessment, while validated, may be subject to response bias. Third, our focus on U.S. institutions limits the generalizability of findings to other academic contexts globally. We deliberately excluded health profession faculty from our analysis due to their distinct clinical practice challenges and COVID-19 related stressors. While this decision enhanced the internal validity of our findings for traditional academic faculty, it limits our ability to speak to the experiences of medical educators and clinical faculty. This will be examined in future analyses but was outside of the current scope. The response rate to our survey introduces potential selection bias, as faculty experiencing severe anxiety might be either more likely to participate due to personal relevance or less likely due to avoidance behaviors. This can also be seen in the higher proportion of tenured full professor faculty responding to the survey, who we noted to experience lower levels of anxiety. Additionally, the timing of our data collection (June-December 2024) may reflect specific temporal stressors in the academic calendar.

Our study did not include analyses of demographic patterns. We excluded demographic covariates aside from gender for two primary reasons: First, our investigation was focused on the interaction between structural and social factors on anxiety among academic faculty and adding demographic factors would change the central research question (e.g., does race or gender explain differences in anxiety levels above and beyond academic discipline). Although our sample had some demographic diversity, such questions would be better answered in a sample with focused recruitment strategies to ensure good representation among the demographic subgroups of interest. Second, demographic covariates added to our analyses would potentially yield uninterpretable results. For example, only age and income were found to have significant relations to other predictors in the sample. If added as covariates, we anticipated problems with multicollinearity (higher ranked professors are older and paid more) and inference (conceptually rank and age are expected to be related). In a review of the literature on academics and mental health, few studies examined demographic differences in mental health among academics, and none used study designs explicitly investigating causal factors related to social identity or demographics and results were inconclusive due to design issues (Urbina-Garcia, 2020). Future studies should explore study designs that can tease apart demographic differences in anxiety concerns among academics.

Another limitation lies in our use of unvalidated measures, with the exception of the GAD-7, in our survey. In the current study, we used face valid dichotomous items (academic parent, close relationships) to understand social supports that were hypothesized to impact anxiety among academics. Although established measures would have been preferred, we are unaware of scale development that focuses on academic experiences of faculty. Future work to develop and refine measures for assessment of constructs that matter to faculty experiences would add to robustness and create opportunities for institutional policy and intervention

Our results suggest several practical implications for higher education institutions. First, universities should consider developing targeted support programs for early-career faculty that explicitly acknowledge and address anxiety. Second, institutions might benefit from fostering stronger community connections, particularly for faculty without academic family backgrounds. Third, the success of HBCU/HSI models in leveraging family support suggests valuable lessons for other institutions in building supportive academic communities. Future research should examine longitudinal changes in faculty anxiety levels, particularly through critical career transitions. Investigation of specific institutional support programs and their effectiveness could provide practical guidance for universities.

Cross-cultural studies comparing faculty anxiety across different national academic systems would enhance the global applicability of these findings.

This study comes at a crucial time when academia faces increasing pressure to address mental health concerns while maintaining academic excellence. By identifying specific moderators of faculty anxiety, our study provides evidence-based guidance for institutions seeking to support faculty well-being while preserving the rigorous standards of academic achievement.

## Declarations

### Ethics approval and consent to participate

This study was approved by the Howard University Institutional Review Board. Informed consent was obtained from all participants. The study was conducted in accordance with relevant guidelines and regulations.

### Availability of data and material

Due to the IRB stipulations, the collected data used for this study cannot be shared or made publicly accessible, however, are available from the corresponding author on reasonable request.

## Authors’ contributions

AA and MH -- Designed the study AA and PM -- Collected the data

NT -- Conducted the statistical analysis

AA, PM, NT, MH -- contributed to writing the paper

## Data Availability

“Due to the IRB stipulations, the collected data used for this study cannot be shared or made publicly accessible, however, are available from the corresponding author on reasonable request.”

## Acknowledgements

N/A

## Clinical trial number

Not applicable.

